# Genome-wide association study identifies multiple HLA loci for sarcoidosis susceptibility

**DOI:** 10.1101/2022.10.13.22281070

**Authors:** SY Liao, S Jacobson, NY Hamzeh, DA Culver, BQ Barkes, P Mroz, K Macphail, K Pacheco, DC Patel, YS Wasfi, LL Koth, CD Langefeld, S Leach, E White, C Montgomery, LA Maier, TE Fingerlin, GRADs investigators

## Abstract

Sarcoidosis is a complex systemic disease. Our study aimed to 1) identify novel alleles associated with sarcoidosis susceptibility; 2) provide an in-depth evaluation of HLA alleles and sarcoidosis susceptibility; 3) integrate genetic and transcription data to identify risk loci that may more directly impact disease pathogenesis.

We report a genome-wide association study of 1,335 sarcoidosis cases and 1,264 controls of European descent (EA) and investigate associated alleles in a study of African Americans (AA: 1,487 cases and 1,504 controls). The EA cohort was recruited from National Jewish Health, Cleveland Clinic, University of California San Francisco, and Genomic Research in Alpha-1 Antitrypsin Deficiency and Sarcoidosis. The AA cohort was from a previous study with subjects enrolled from multiple United States sites. HLA alleles were imputed and tested for association with sarcoidosis susceptibility. Expression quantitative locus and colocalization analysis were performed using a subset of subjects with transcriptome data.

49 SNPs in *HLA-DRA, -DRB9, -DRB5, -DQA1*, and *BRD2* genes were significantly associated with sarcoidosis susceptibility in EA. Among them, rs3129888 was also a risk variant for sarcoidosis in AA. Classical HLA alleles DRB1*0101, DQA1*0101, and DQB1*0501, which are highly correlated, were also associated with sarcoidosis. rs3135287 near *HLA-DRA* was associated with *HLA-DRA* expression in peripheral blood mononuclear cells and bronchoalveolar lavage.

In summary, we identified several novel SNPs and three HLA alleles associated with sarcoidosis susceptibility in the largest EA population evaluated to date using an integrative analysis of genetics and transcriptomics. We also replicated our findings in an AA population.

## Introduction

Sarcoidosis is a complex systemic disease affecting between 45-300/100,000 persons in the United States[1]. While environmental and genetic susceptibility factors have been associated with sarcoidosis[2-4], the driving risk factors for disease are still largely undefined. Previous genome-wide association studies (GWAS) have consistently implicated the HLA region as harboring genetic risk loci based on single nucleotide polymorphisms (SNPs)[5-12], while candidate gene studies have implicated specific HLA alleles such as DRB1*1101, DRB1*1501, and DQB1*0602[13, 14]. The strong role of the HLA region for sarcoidosis disease risk is consistent with exposure and immune-based associations observed in patients, as well as the heterogeneous disease course.

Most sarcoidosis genetic studies have focused on genotype data, limiting understanding of potential functional genetic features associated with disease status. Studying the impact of risk alleles on gene expression can provide the first step in understanding their potential biological impact[15]. This is especially true for complex diseases since the majority of genetic variants robustly associated with these diseases fall in non-coding regions of the genome[16]. While the function of non-coding regions was unknown in the past, there is substantial evidence that many of them influence disease risk through regulatory effects, including those that impact gene expression[15]. Hence, integrating genetic and transcriptomic data may identify loci with a more direct or functional effect on disease pathogenesis.

To identify genetic risk factors for disease and study their effects on gene expression, we conducted a genome-wide association study in European American (EA) population that includes follow-up of risk alleles on gene expression in peripheral blood as well as lung cells. We confirm the strong role of the HLA region in sarcoidosis risk and demonstrate both novel SNPs and classical HLA alleles associated with disease and influencing HLA expression. We replicated some of these findings in a large African American (AA) population.

## Materials and Methods

### Study design and population

This GWAS used a two-phase approach to identify genome-wide significant SNPs associated with sarcoidosis; additional details are present in Supplemental Methods. DNA samples from sarcoidosis cases and controls were obtained from National Jewish Health (NJH), University of California, San Francisco, Genomic Research in Alpha-1 Antitrypsin Deficiency and Sarcoidosis (GRADS) consortium [17], and Cleveland Clinic Foundation. Sarcoidosis case definition was based on the ATS/ERS/WASOG statement[18]. After quality control (see below), 818 cases and 981 controls (Phase 1) and 517 cases and 283 controls (Phase 2), all self-reported EA ancestry (**Table** 1 and **Figure 1**), reflected the majority of the race/ethnicities seen in our clinics. A subset of study participants had peripheral blood mononuclear cell (PBMC) and/or bronchoalveolar lavage (BAL) cell RNA sequencing data available through GRADS study[17, 19]. We tested significant SNPs identified in our EA two-phase approach in an AA study population. The AA GWAS summary statistics were obtained from a published study[5] with updated imputations since publication of those data.

**Figure 1.**
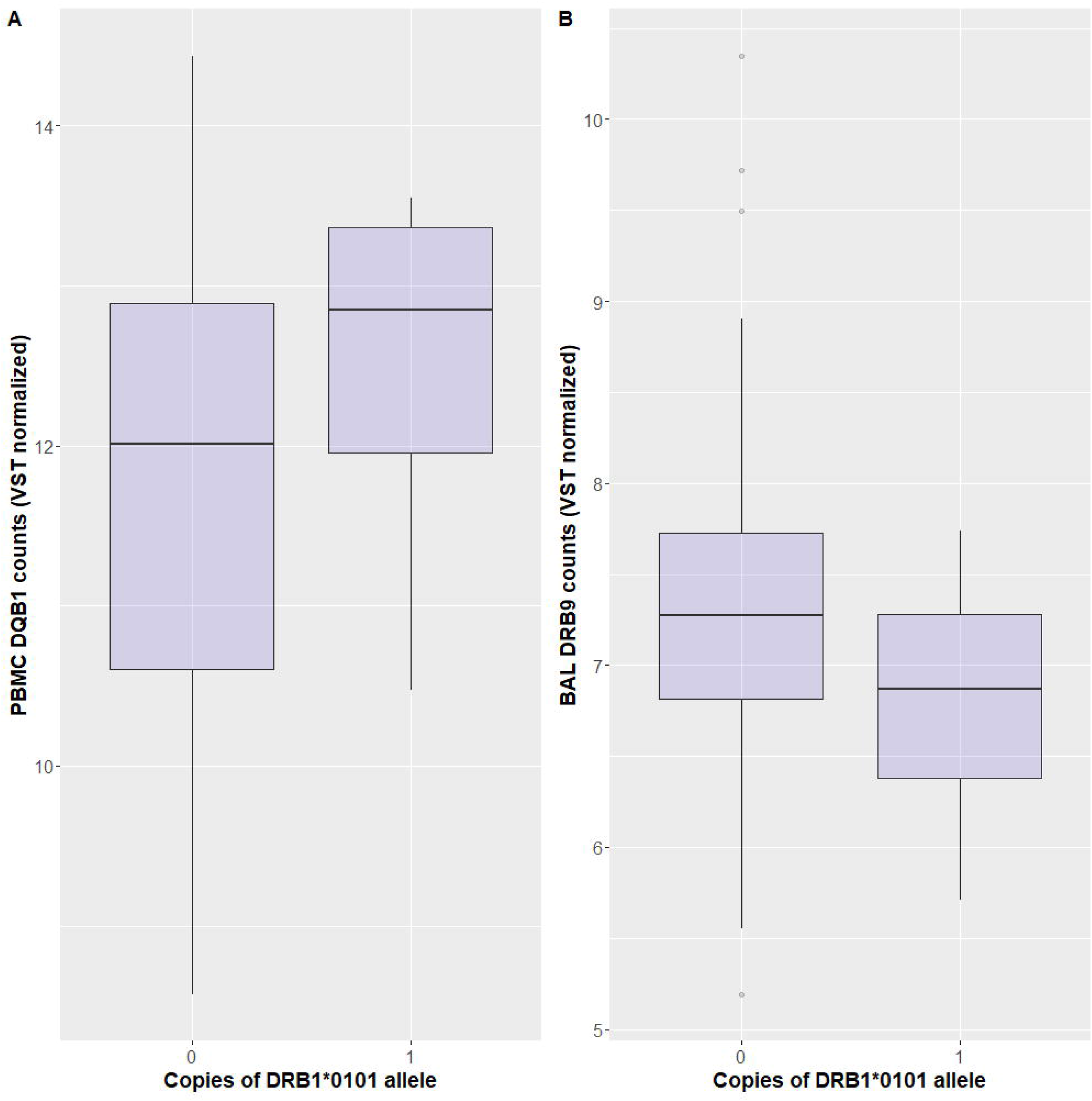
CONSORT flow chart for the GWAS samples. This figure outlines the exclusion of the subject in Phase 1 and Phase 2 respectively.

### Genome-wide genotyping

DNA was extracted from whole blood using the PAXgene Blood DNA kit. Genotyping was performed using the Illumina HumanOmini 2.5 BeadChip to interrogate ∼ 2.4 million markers. The markers were derived from the 1000 Genomes Project[21], including all three HapMap phases, 19K SNPs across the MHC, and over 41K non-synonymous SNPs. Genotyping was conducted at Hudson Alpha Biotechnology Institute (Huntsville, AL https://hudsonalpha.org/).

### Genotype quality control

We used 1000 genomes data[21] to infer ancestry-informative principal components (PCs), which were projected onto cases and controls. We prioritized SNPs with minor allele frequency (MAF)>0.03 and Hardy-Weinberg Equilibrium p>0.001 in cases and controls evaluated separately and <10% missing data.

### Imputation of additional genotypes and HLA variants for the EA population

We imputed genotypes using combined case and control discovery samples for all 1000 genomes SNPs. We imputed classical HLA alleles using R package HLA Genotype Imputation with Attribute Bagging (HIBAG)[27].

### RNA sequencing and quality control

Total RNA was extracted, and RNA-sequencing was conducted as outlined[19]. We followed a similar quality control procedure for both PBMC and BAL samples and removed RNA samples with an unmapped read rate >20% and mitochondrial read rate >0%. We removed outlier samples through PC analysis; the EA individuals who also had gene expression data available were included in the expression quantitative trait analyses.

### Statistical analysis

#### Single-SNP association test and meta-analysis

We tested for association between each SNP and sarcoidosis using Snptest (v2)[28] as described previously[29]. To obtain an overall measure of association with sarcoidosis, we performed a meta-analysis of Phase 1 and Phase 2 using summary statistic data and the weighted inverse normal method[30] as implemented in the software METAL[31]. Genome-wide significance was defined as meta-analysis p<5×10^−8^. Genome-wide significant SNPs identified in our EA population were tested in the AA population. Statistical significance for these SNPs was defined as p<0.05/(number of significant SNPs in EA). We compared our GWAS results to previously identified loci in other studies including SNPs in *ANXA11* (rs1049550, rs1953600, rs2573346, rs2784773)[32], *RAB23* (rs1040461)[9], *C100RF67* (rs1398024)[33], *OS9* (rs1050045)[8], *CCDC88B* (rs479777)[7], and *NOTCH4* (rs715299)[5]. Statistical significance for these *a priori* SNPs was defined as p<0.0056(0.05/9 SNPs).

#### Classic HLA alleles analysis

We used logistic regression models to test for association between dosage of each imputed HLA allele and sarcoidosis. Given strong *a priori* associations with the HLA region, we used p<0.00011(0.05/448 HLA alleles tested) as statistical significance. We compared our HLA results to previously identified HLA alleles in other studies, including DRB1*1101, DRB1*1501, and DQB1*0602[13, 14]. We used a p=0.016(0.05/3 alleles) to determine statistical significance for *a priori* alleles.

#### Conditional Models

To assess the independence of single-SNP effects from HLA risk alleles, we computed multivariable logistic regression models where HLA risk alleles were included as covariates in the model, and each SNP, one at a time, was tested for association (i.e., association adjusted for HLA risk alleles).

#### Expression quantitative locus (eQTL) and colocalization analysis

For those sarcoidosis cases with gene expression data from GRADS, we performed colocalization analysis using eCAVIAR[34] to identify variants with evidence for colocalization of disease and *cis* eQTL associations. The algorithm estimates the posterior probability that the same variant is casual in both GWAS and eQTL studies while accounting for linkage disequilibrium (LD). We included all SNPs within the defined gene boundary of the significant SNP in the analysis (+/-25K base pairs) and tested for association between those SNPs and gene expression. The threshold for significance was set as a colocalization posterior probability (CLPP)>0.001, as suggested by eCAVIAR[34]. The same approach was applied to imputed HLA alleles. We tested for association between HLA alleles and gene expression in two tissues (BAL and PBMC). In addition, using GRADS data, we conducted a comprehensive cis-eQTL search using the publicly available database, Genotype-Tissue Expression (GTEx)[36, 37]. The GTEx project was supported by the Common Fund of the Office of the Director of National Institutes of Health, NCI, NHGRI, NHLBI, NIDA, NIMH, and NINDS. The data used for analyses described in this manuscript were obtained from the GTEx Portal on 05/31/2022.

## Results

### Descriptive analysis of GWAS in EA

We enrolled 3,141 subjects genotyped on the Illumina array. After quality control (**Figure 1**), we included n=2,599 subjects (1,335 cases and 1,264 controls) in the analysis (**Table 1**); 818 cases (49% female/51% male) and 981 controls (37% female/63% male) in Phase 1 and 517 cases (53% female/47% male) and 283 controls (71% male/29 % female) in Phase 2. For eQTL analyses, paired genotype-transcription data was available for 136 genotype-BAL transcription and 193 genotype-PBMC transcription samples.

**Table 1:** Sample size and sex for Phase 1 and Phase 2*

|  | All (N= 2599) |  | Phase 1 (N=1799) |  | Phase 2 (N=800) |  |
| --- | --- | --- | --- | --- | --- | --- |
| Sex | Sarcoidosis<br>(n= 1335) | Control<br>(n=1264) | Sarcoidosis<br>(n=818) | Control<br>(n=981) | Sarcoidosis<br>(n=517) | Control<br>(n=283) |
| Female, n (%) | 671 (50 %) | 565 (45%) | 399 (49%) | 364 (37%) | 272 (53%) | 201 (71%) |
| Male, n (%) | 664 (50%) | 699 (55%) | 419 (51%) | 617 (63%) | 245 (47%) | 82 (29%) |

\*Phase 2 DNA samples became available after the Phase 1 group had been genotyped (see Materials and Methods).

### GWAS identifies HLA region associations with sarcoidosis

The meta-analysis of Phase 1 and 2 data identified 49 SNPs reaching GWAS significance. The *r*^*2*^ LD plot of those SNPs and the top seven SNPs representing all significant SNPs (*r*^*2*^>0.70) are shown in **Figure S1**. Those top seven SNPs were rs9269233, rs9271346, rs35656642, rs28589559, rs9276935, rs3129888, and rs71549283 (**Table 2**), located across *HLA-DRA, - DRB9, -DRB5, -DQA1*, and *BRD2 on* chromosome 6. The remaining significant SNPs are shown in **Table S1**. With the much-reduced Phase 2 sample size compared to Phase 1, the Phase 2 p-value is not nominally significant, although effect sizes (odds ratios) were comparable, and both contribute proportionally to meta-analysis p-values. The Manhattan plot for SNP associations **is shown in Figure 2A**, and **Figure 2B** shows the locus-specific plot for all significant SNPs highlighting the seven top SNPs. For the seven top SNPs, we used a stepwise approach to adjust for other SNPs (**Table S2**), and found rs9269233, rs9276935, rs28589559, and rs3129888 still nominally significant after adjustment (all p<0.01). Other top SNPs with p<5 × 10^−5^ are shown in **Table S3**. In the AA cohort, rs3129888 was also significantly associated with increased risk of sarcoidosis (**Table 2**). When we compared our results to previously-identified loci from other GWAS studies, we found SNPs in *ANXA11* were nominally significant (**Table S4**) in the meta-analysis (p<0.0056).

**Table 2.**
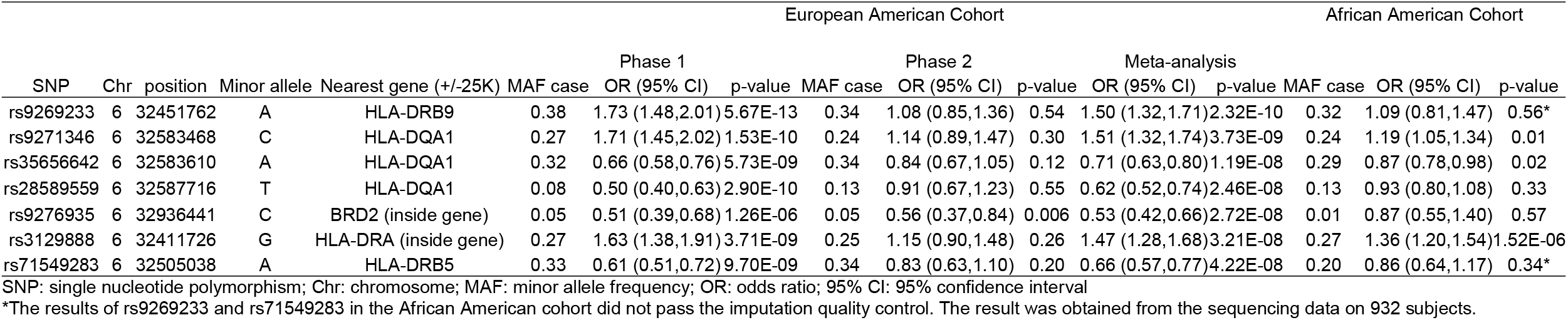
Summary of top 7 genome-wide significant SNPs of the genome-wide association study of sarcoidosis (p-value<5×10^−8^)

**Figure 2.**
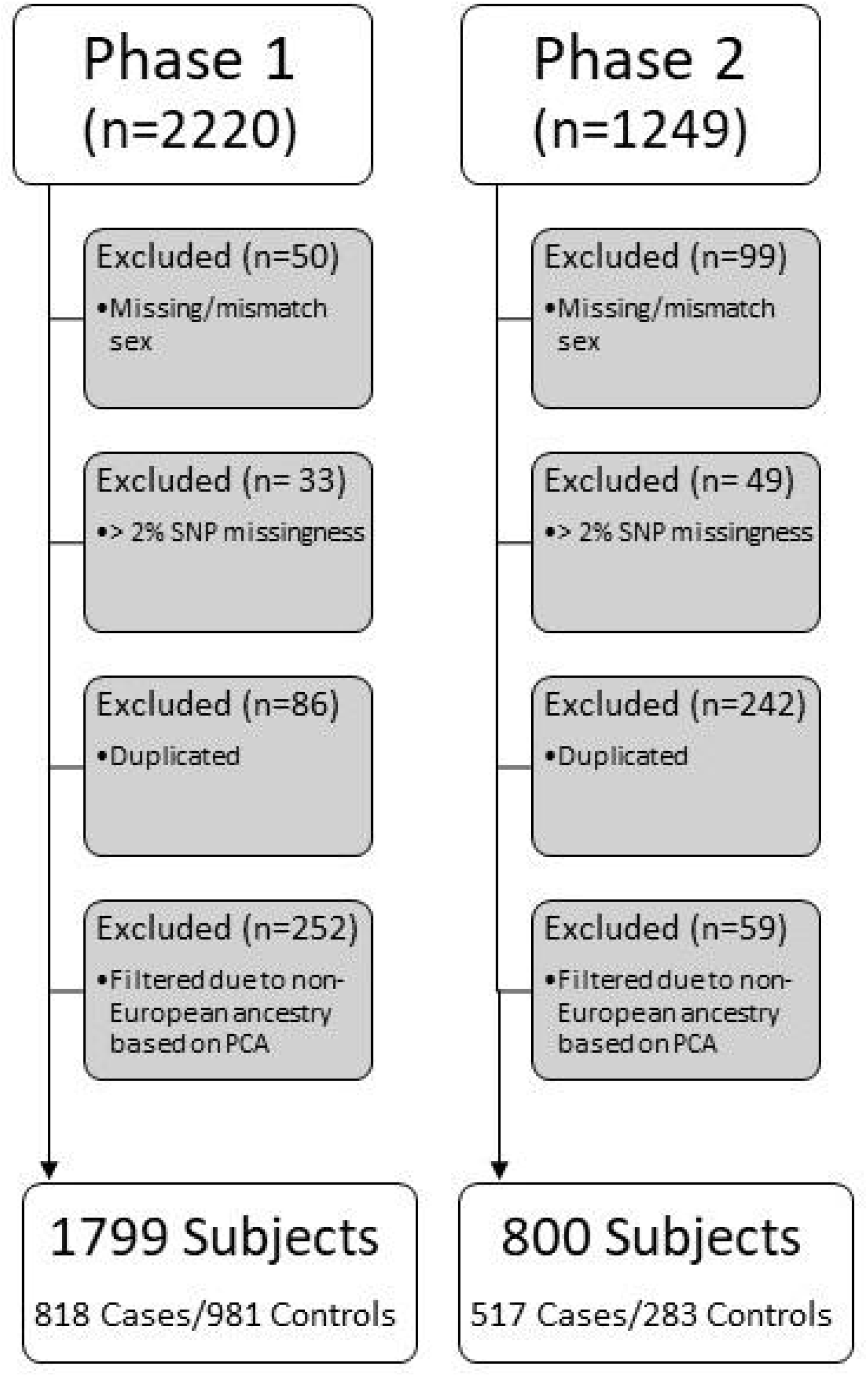
2A) Manhattan plot for SNP associations, with significant associations in the HLA region of chromosome 6. 2B) Locus-zoom plot for significant SNPs in chromosome 6 (position: 32381726-32984689); rs9269233 is the most significant SNP and *r*^2^ values with other significant SNPs are low.

### Classic HLA alleles are associated with sarcoidosis

Nine HLA alleles were significantly associated with sarcoidosis susceptibility (p<0.00011, **Table 3 and Table S5**). Three HLA alleles had a p-value<5 × 10^−8^: DRB1*0101, DQA1*0101, and DQB1*0501. All three HLA alleles were highly correlated and protective against sarcoidosis. We found three candidate HLA alleles, DRB1*1101, DRB1*1501, and DQB1*0602, significantly associated with increased risk of sarcoidosis (**Table 3**).

**Table 3.**
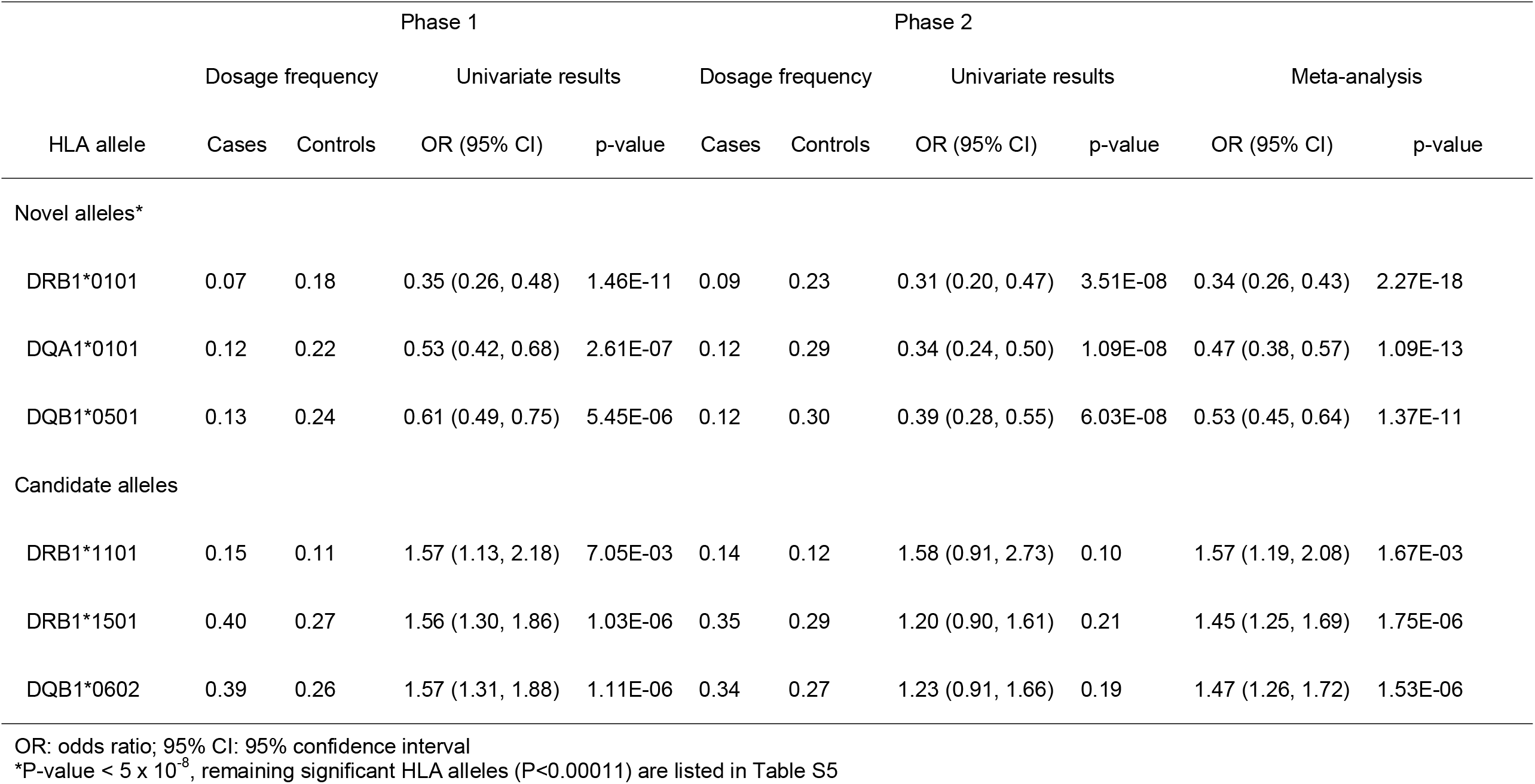
Association between HLA alleles and sarcoidosis.

### GWAS SNPs in HLA are independent of HLA allele associations

Although the p-values were attenuated slightly, each genome-wide significant SNP remained associated with sarcoidosis after adjustment for each of the HLA risk alleles (**Table 4)**. We then adjusted for all three HLA risk alleles, and the effects of each SNP on odds ratios were largely unchanged after adjustment (**Table S6**). Interestingly, there was a significant interaction between rs9271346 and DRB1*0101 (p=0.02, **Table S7**).

**Table 4.**
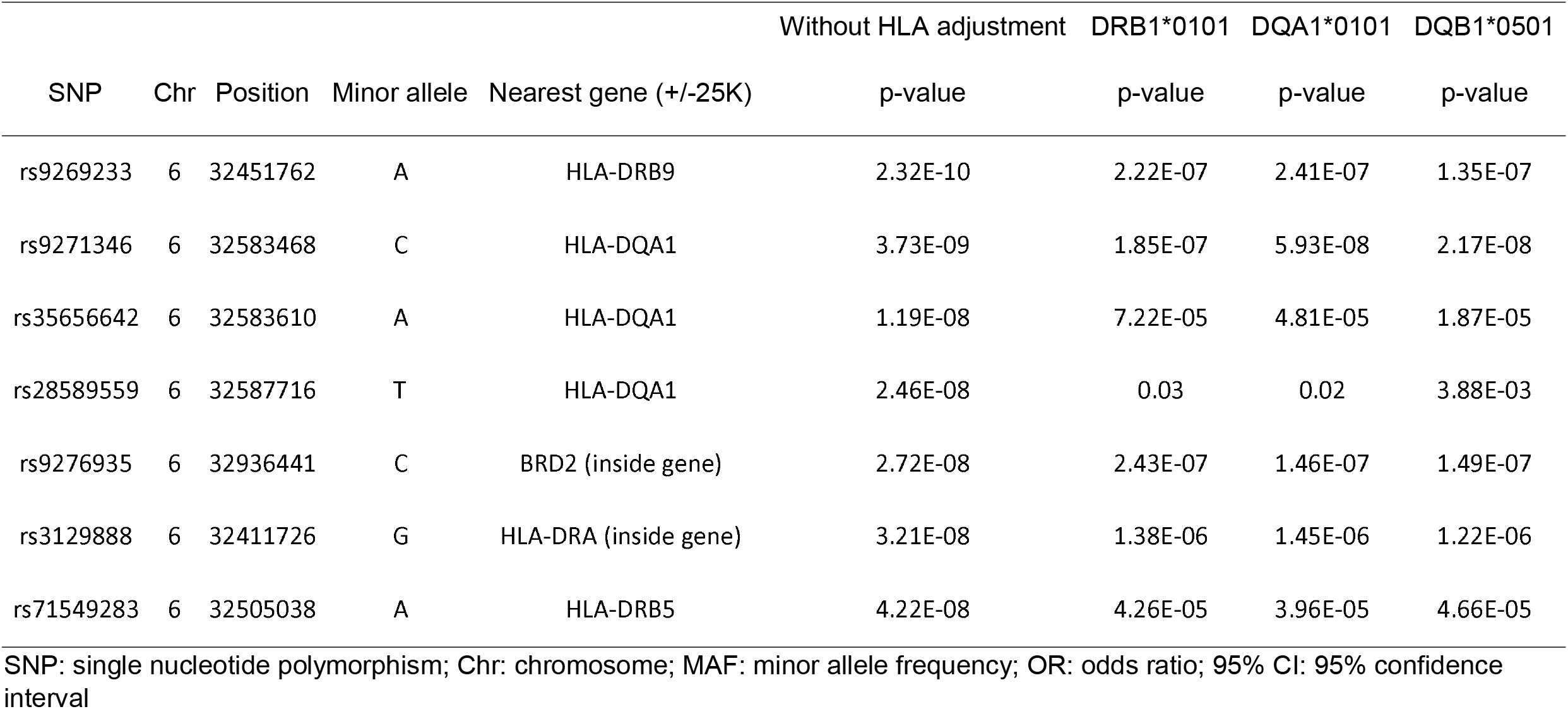
P-values of the three genome-wide significant SNPs adjusted by HLA alleles (one at a time)

### eQTL and colocalization analyses demonstrate an association between SNPs and gene expression

Colocalization analysis was conducted across five genes: *HLA-DRA, -DRB9, -DRB5, -DQA1*, and *BRD2*. We found no significant colocalization when we assumed only one casual SNP in the region. The model assuming two casual SNPs in the region demonstrated that rs3135387 colocalized with both PBMC and BAL cell expression levels of *HLA-DRA* (CLPP=0.003 and 0.002, respectively). Other nearby SNPs, rs3129888, and rs3135390, within the *HLA-DRA* region also showed significant colocalization posterior probability with PBMC gene expression (CLPP=0.002, **Figure S2)**. A comprehensive cis-eQTL search of whole blood and lung from GTEx is shown in **Table 5**. In brief, rs9269233 demonstrated significant eQTL for *HLA-DRB9* expression in whole blood, while rs9271346, rs35656642, and rs28589559 were significant eQTLs for expression of H*LA-DQA1* in whole blood and lung. The colocalization analysis for the three genome-wide significant HLA alleles showed that DRB1*0101 was the most significantly associated with sarcoidosis susceptibility and with expression levels in PBMC and BAL among sarcoidosis patients for *DRB1, DQA1, DQB1*, and *DRB9* (all CLPPs>0.001). The DRB1*0101 risk allele was positively associated with PBMC *DQB1* gene expression (p=0.03, **Figure 3A**) and negatively associated with BAL *DRB9* gene expression (p=0.03, **Figure 3B**).

**Table 5.** Genotype-Tissue Expression Portal (GTEx) *Cis*-eQTL in lung and whole blood for associated SNPs.

| SNP | Nearest gene (+/- 25K) | Common for Lung and Whole blood | *Unique for Lung | *Unique for Whole blood |
| --- | --- | --- | --- | --- |
| rs9269233 | HLA-DRB9 | HLA-DQA2, HLA-DQB1, HLA-DQB1-AS1, HLA-DRB1, HLA-DRB5, HLA-DRB6 | LY6G5C, STK19B | C4A, CYP21A2, HLA-DQB2, HLA-DRB9 |
| rs9271346 | HLA-DQA1 | CYP21A2, HLA-DQA1, HLA-DQA2, HLA-DQB1, HLA-DQB1-AS1, HLA-DQB2, HLA-DRB1, HLA-DRB5, HLA-DRB6, HLA-DRB9 | HCG23, XXbac-BPG154L12.4 | LY6G5B, TNXB |
| rs35656642 | HLA-DQA1 | HLA-DQA1, HLA-DQA2, HLA-DRB5, HLA-DRB6, LY6G5B | HCG23, LY6G5C, XXbac-BPG154L12.4 | CYP21A1P, HLA-DRB1 |
| rs28589559 | HLA-DQA1 | HLA-DQA1, HLA-DQA2, HLA-DQB2, HLA-DRB6, HLA-DRB9 | HLA-DOB, NOTCH4, TAP2 | HLA-DQB1, PBX2 |
| rs3129888 | HLA-DRA (inside gene) | C4A, CYP21A2, HLA-DQA2, HLA-DQB1, HLA-DQB1-AS1, HLA-DRB5, HLA-DRB6, HLA-DRB9, STK19B | HCG23, HLA-DQA1, HLA-DRB1, XXbac-BPG154L12.4 | HLA-DQB2 |

\*Unique gene with its gene expression affected by the specific SNPs only in the specific tissue. For example. rs9269233 only affects *STK19B* gene expression in the lung but not in the whole blood.
rs9276935 were not eQTLs in lung or whole blood tissue; rs71549283 was not found in the GTEx database as of 5/31/2022

**Figure 3.**
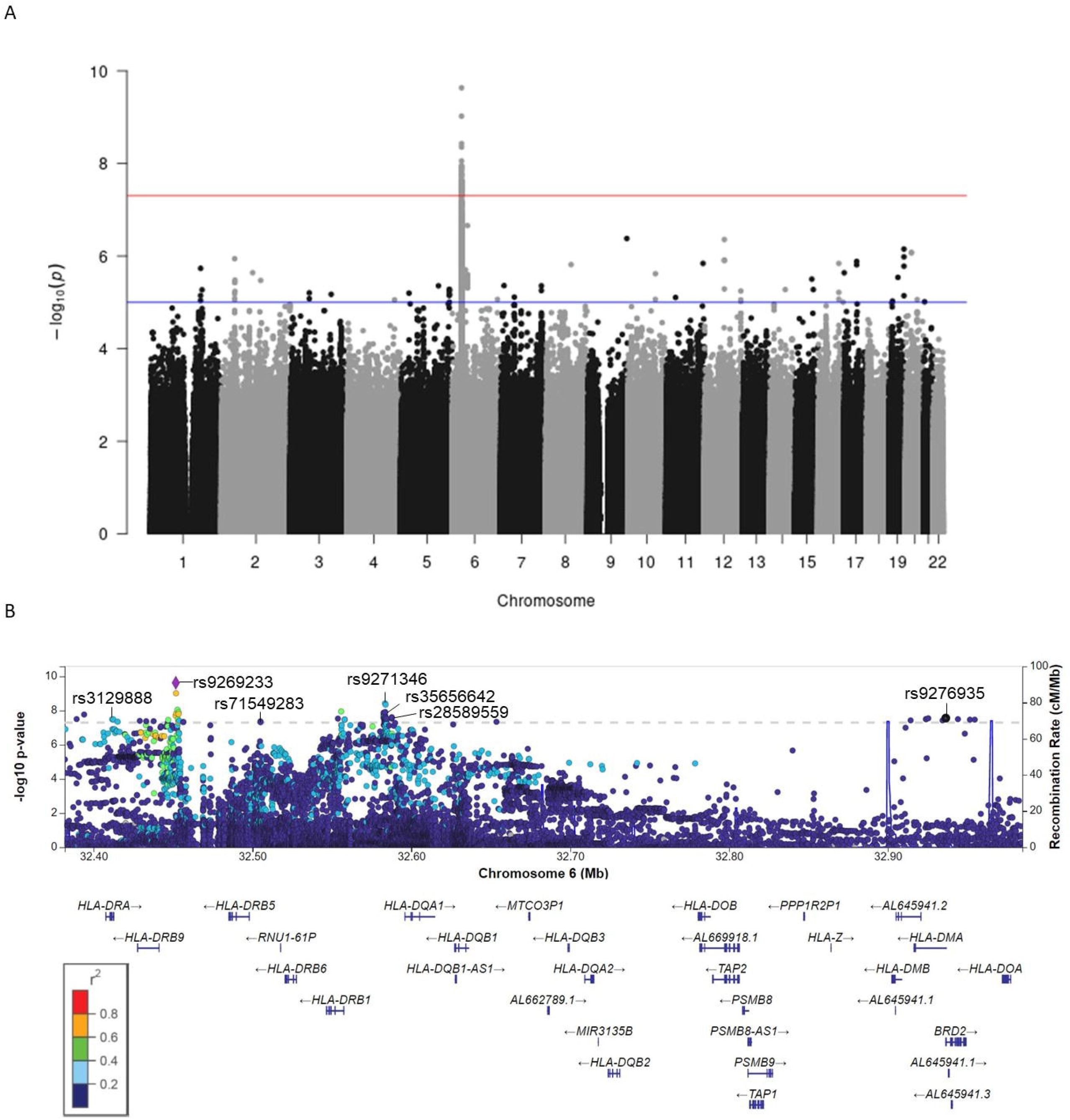
A) PBMC DQB1 gene expression (variance stabilizing transformation [VST] normalized counts) and presence/absence of DRB1*0101 allele. Individuals with one DRB1*0101 allele have higher PBMC DQB1 gene expression compared to those with none. B) BAL DRB9 gene expression (VST normalized counts) and presence/absence of DRB1*0101 allele. Individuals with one DRB1*0101 allele have higher BAL DRB9 gene expression.

## Discussion

We identified 49 SNPs associated with sarcoidosis in this largest EA GWAS of sarcoidosis; one of these SNPs, rs3129888, was also associated with sarcoidosis in an AA GWAS. All SNPs are on Chromosome 6 in the well-known HLA risk region. While we evaluated previously identified GWAS SNPs *a priori* and found an association with *ANXA11*, we found no other SNPs outside the HLA region associated with sarcoidosis. Using colocalization and eQTL analysis, we found rs3135387 colocalizes with PBMC and BAL gene expression of *HLA-DRA* in our EA population. HLA DRB1*0101, DQA1*0101, and DQB1*05*01 were significantly associated with sarcoidosis, while DRB1*0101 was also associated with expression of PBMC *HLA-DQB1* and BAL *HLA-DRB9*. These results suggest that in sarcoidosis, the HLA region is likely functional, impacting gene expression and disease development.

The most significant SNP associated with sarcoidosis in our study, rs9269233, is between *HLA-DRB9* and *HLA-DRB5*. This association with rs9269233 was only modestly attenuated after adjustment for sarcoidosis-related HLA alleles (adjusted p=2.15×10^−7^), suggesting that rs9269233 is an independent risk allele for sarcoidosis in the region. The A allele of this SNP showed increased risk of sarcoidosis, has not been reported in other studies and was not significant in AA cohort. Potentially this may indicate that sarcoidosis pathogenesis differs between EA and AA, and in fact, other studies have found different risk variants in EA and AA subjects[38]. In a previous GWAS[38], rs1964995, also located between *HLA-DRB9* and *HLA-DRB5* (*r*^*2*^ with rs9269233=0.53), showed a protective effect in non-Lofgren sarcoidosis vs. controls in a white Swedish cohort. Interestingly, rs1964995 increases the risk of rheumatoid arthritis (RA) in AA, although not in EA[39]. rs9269233 has been found to be an eQTL for *HLA-DRB9* gene expression in whole blood in our analysis using the GTEx database. In addition to rs9269233, the presence of at least one DRB1*0101 allele was significantly associated with *HLA-DRB9* gene expression in BAL cells. *HLA-DRB9* is a pseudogene that is transcribed into RNA but does not encode proteins. However, pseudogenes can regulate other protein-coding genes[40]; our findings suggest this may be the case with DRB9 in our population. In previous studies, *HLA-DRB* loci were found to group into five major haplogroups (DR1, DR8, DR51, DR52, and DR53), which differ by alleles at functional DRB genes (DRB3, DRB4, or DRB5) and DRB pseudogenes (DRB2, DRB6, DRB7, DRB8, or DRB9)[41]. The HLA-DRs are associated with various diseases, including sarcoidosis[42], which is usually thought to have implications for antigen presentation. Our data suggests that these associations may reflect gene expression regulation that is not currently known to be functional but may impact immune responses in sarcoidosis.

Two other significant SNPs associated with sarcoidosis, rs9271346, and rs35656642, are both near *HLA-DQA1* (<25K base pairs). rs9271346 was nominally significant in AA and EA in a previous study[5], with the same allele associated with risk in each, although it did not reach genome-wide significance (p=0.007 in AA and 8.63×10^−6^ in EA). rs35656642 is a novel risk variant for sarcoidosis that has not been described previously, and we found the same allele associated with sarcoidosis in AA. Both SNPs are also eQTLs in lung tissue and whole blood based on publicly available datasets, although we did not find associations in lung BAL cells in our study. A study found rs2187668 near gene *HLA-DQA1* significantly associated with Lofgren’s syndrome[38], while another reported three SNPs (rs28609302, rs9273113, rs9272594) in this region associated with ocular sarcoidosis vs. controls in US AA(2 × 10-7 to 6 × 10^−6^)[43]. Of note, a subpopulation from the AA cohort was used in our study. We found low LD between those previously reported SNPs and the SNPs in our EA population(absolute *r*^*2*^ 0.06-0.58), indicating that the findings in our EA population are unlikely simple replications of previously-identified SNPs.

Except for rs3129888 (p=1.52×10^−6^), significant SNPs in EA were not significantly associated with sarcoidosis in AA. The rs3129888 G allele, located in an *HLA-DRA* intron, increased sarcoidosis risk. This finding is consistent with a study on Lofgren’s syndrome in Sweden, Germany, and a US AA population[38]. Our US EA samples are distinct from the European cohort, but our AA population largely overlaps with the US AA population[38].

We found three HLA alleles significantly associated with sarcoidosis, all of which are highly correlated with each other. DRB1*0101 and DQB1*0501 showed a protective effect on sarcoidosis risk, consistent with a previous study from the UK, Netherlands, and Japan[44]. In this previous study, DRB1*0101 was protective against lung-predominant sarcoidosis, Lofgren’s syndrome, and uveitis. The sarcoidosis patients in our study included all subtypes of disease. When we searched the database from another large study using 13,835 EA individuals from five US sites of the Electronic Medical Records and Genomics (eMERGE) network[45] (using the International Classification of Diseases code as the definition for diseases), DRB1*0101, DQB1*0501, and DQA1*0101 were all protective for sarcoidosis but increased risk of RA (**Table S8**). These opposite genetic effects are consistent with a study of pleiotropy between sarcoidosis and RA, which demonstrates higher RA polygenic risk score associated with a protective effect of sarcoidosis[46], and another epidemiologic study demonstrating a lower prevalence of RA in a British sarcoidosis cohort vs. the general population[47]. While RA and sarcoidosis are both inflammatory diseases, this may imply that their disease pathogenesis is distinct and drivers of one disease protect from development of the other. Each of the previously-reported sarcoidosis HLA risk alleles, DRB1*1101, DRB1*1501, and DQB1*0602, showed a nominal increase in the risk of sarcoidosis in our study. DRB1*1101 allele was previously associated with increased risk of sarcoidosis in AA and European descent individuals[14]. DRB1*1501, which is in high LD with DQB1*0602, has been associated with increased risk of severe pulmonary sarcoidosis in individuals of European descent[13]. Of note, these HLA alleles were not the strongest risk alleles in our study, and this might be due to sub-populations (e.g., race/ethnicity) or varied phenotypes in our cohort compared to others. For example, our cohort is likely a mixture of different sarcoidosis phenotypes (e.g., cardiac, ocular, cutaneous, etc.) as we did not restrict enrollment to specific phenotypes. Future GWAS studies would benefit from focusing on specific sarcoidosis phenotypes or those with specific organ involvement to explore genetic drivers of sarcoidosis manifestations, including neurological, cardiac, or specific pulmonary phenotypes of sarcoidosis.

There are limitations to our study. First, we included a heterogeneous population of sarcoidosis patients we various phenotypes; this could reduce our study’s power to identify novel SNPs versus European studies focused on Lofgren’s syndrome. Regardless, we found HLA associations linked to other sarcoidosis phenotypes in previous studies, like DRB1*1101 and *1501. Second, we have gene expression data available on only a subset of participants for the eQTL analyses, impacting power to identify other eQTLs in our study. To help mitigate this limitation, we used the GTEx database to enhance our evaluation of associations between SNPs and gene expression.

In summary, our findings provide convincing evidence that HLA alleles are an important contributor to risk of sarcoidosis not only in our EA population but also in an AA population with genotyping already available. In addition to traditional GWAS SNPs and imputed HLA variants, we also explored how these genotypes are associated with gene expression in both PBMC and BAL cells using integrated analysis and demonstrated showed potential gene expressions impacted by these risk variants. Our future studies will explore these potential variants/genes and their mechanistic implications.

## Data Availability

All data produced in the present study are available upon reasonable request to the authors

## Acknowledgments

We would like to thank all the participants of this study, as well as for the administrative support that we received from NJH and research coordination assistance from Christina Riley. We also appreciate the grant support from National Institutes of Health Grants U01 HL112707, U01 HL112694, U01 HL112695, U01 HL112696, U01 HL112702, U01 HL112708, U01 HL112711, U01 HL112712, UL1 RR029882, UL1 RR025780, R01 HL110883, R01 HL114587, R01HL114587; Clinical and Translational Science Institute Grant U54 9 UL1 TR000005; and Centers for Disease Control and Prevention National Mesothelioma Virtual Bank for Translational Research Grant 5 U24 OH009077; and Foundation for Sarcoidosis Research (FSR).

## Declaration of interests

All authors report no conflicts of interest related to this work

## Notes

### Competing Interest Statement

The authors have declared no competing interest.

### Funding Statement

This study was funded by National Institutes of Health Grants U01 HL112707, U01 HL112694, U01 HL112695, U01 HL112696, U01 HL112702, U01 HL112708, U01 HL112711, U01 HL112712, UL1 RR029882, UL1 RR025780, R01 HL110883, R01 HL114587, R01HL114587; Clinical and Translational Science Institute Grant U54 9 UL1 TR000005; and Centers for Disease Control and Prevention National Mesothelioma Virtual Bank for Translational Research Grant 5 U24 OH009077; and Foundation for Sarcoidosis Research (FSR).

### Author Declarations

Institutional Review Board at National Jewish Health (HS2765) gave ethical approval for this work. Institutional review board through Genomic Research in Alpha-1 Antitrypsin Deficiency and Sarcoidosis (GRADS) (HS2780) gave ethical approval for this work.

