## Supplemental Table for "Genome-wide association study identifies multiple HLA loci for sarcoidosis susceptibility"

| **Table S1**. Summary of additional genome-wide significant SNPs of the genome-wide association study of sarcoidosis (p-value<5x10^-8^) | | | | | | | | | | | | | | | | | | | |
| --- | --- | --- | --- | --- | --- | --- | --- | --- | --- | --- | --- | --- | --- | --- | --- | --- | --- | --- | --- |
|  | | | | |  | European American Cohort | | | | | | | | | |  | African American Cohort | | |
|  |  |  |  |  |  | Phase 1 | | |  | Phase 2 | | |  | Meta-analysis | |  |  |  |  |
| SNP | Chr | position | Minor allele | Nearest gene (+/-25K) |  | MAF case | OR (95% CI) | p-value |  | MAF case | OR (95% CI) | p-value |  | OR (95% CI) | p-value |  | MAF case | OR (95% CI) | p-value |
| rs2395202 | 6 | 32451906 | T | HLA-DRB9 |  | 0.44 | 1.68 (1.46,1.94) | 5.99E-13 |  | 0.4 | 1.03 (0.82,1.28) | 0.82 |  | 1.45 (1.29,1.64) | 9.54E-10 |  | 0.36 | 0.99 (0.71,1.39) | 0.96 |
| rs9271349 | 6 | 32583830 | A | HLA-DQA1 |  | 0.27 | 1.71 (1.45,2.02) | 1.98E-10 |  | 0.24 | 1.14 (0.89,1.47) | 0.30 |  | 1.51 (1.31,1.74) | 4.43E-09 |  | 0.24 | 1.19 (1.05,1.34) | 0.01 |
| rs28698533 | 6 | 32452802 | T | HLA-DRB9 |  | 0.42 | 1.65 (1.42,1.93) | 5.18E-11 |  | 0.39 | 1.06 (0.84,1.35) | 0.60 |  | 1.45 (1.28,1.65) | 8.83E-09 |  | 0.45 | 1.41 (0.9,2.21) | 0.13 |
| rs13204736 | 6 | 32582603 | A | HLA-DQA1 |  | 0.27 | 0.64 (0.55,0.73) | 3.74E-10 |  | 0.31 | 0.91 (0.72,1.14) | 0.40 |  | 0.7 (0.62,0.79) | 1.33E-08 |  | 0.31 | 0.99 (0.76,1.29) | 0.94* |
| rs2395206 | 6 | 32452176 | G | HLA-DRB9 |  | 0.46 | 1.63 (1.41,1.88) | 1.23E-11 |  | 0.43 | 1.01 (0.81,1.25) | 0.95 |  | 1.41 (1.25,1.59) | 1.39E-08 |  | 0.44 | 0.96 (0.71,1.3) | 0.79** |
| rs34341095 | 6 | 32583605 | G | HLA-DQA1 |  | 0.32 | 0.66 (0.58,0.76) | 4.66E-09 |  | 0.35 | 0.86 (0.69,1.07) | 0.18 |  | 0.71 (0.63,0.8) | 1.89E-08 |  | 0.31 | 0.87 (0.78,0.97) | 0.01 |
| rs28619202 | 6 | 32451650 | T | HLA-DRB9 |  | 0.43 | 1.63 (1.41,1.89) | 2.88E-11 |  | 0.4 | 1.02 (0.82,1.27) | 0.88 |  | 1.41 (1.25,1.6) | 1.95E-08 |  | 0.37 | 1 (0.71,1.41) | 0.98 |
| rs35186928 | 6 | 32582604 | A | HLA-DQA1 |  | 0.26 | 0.64 (0.55,0.74) | 9.44E-10 |  | 0.31 | 0.9 (0.72,1.13) | 0.37 |  | 0.71 (0.62,0.8) | 2.32E-08 |  | 0.26 | 0.94 (0.73,1.23) | 0.66 |
| rs34656207 | 6 | 32582601 | T | HLA-DQA1 |  | 0.26 | 0.64 (0.55,0.74) | 9.44E-10 |  | 0.31 | 0.9 (0.72,1.13) | 0.37 |  | 0.71 (0.62,0.8) | 2.32E-08 |  | 0.26 | 0.94 (0.73,1.23) | 0.66 |
| rs34599306 | 6 | 32583557 | G | HLA-DQA1 |  | 0.32 | 0.66 (0.58,0.76) | 5.57E-09 |  | 0.35 | 0.86 (0.69,1.08) | 0.19 |  | 0.71 (0.63,0.8) | 2.46E-08 |  | 0.31 | 0.87 (0.78,0.97) | 0.01 |
| rs28707527 | 6 | 32587717 | A | HLA-DQA1 |  | 0.08 | 0.5 (0.4,0.63) | 2.9E-10 |  | 0.13 | 0.91 (0.67,1.23) | 0.55 |  | 0.62 (0.52,0.74) | 2.46E-08 |  | 0.13 | 0.93 (0.8,1.08) | 0.33 |
| rs36124427 | 6 | 32583677 | C | HLA-DQA1 |  | 0.32 | 0.66 (0.58,0.76) | 5.09E-09 |  | 0.35 | 0.86 (0.69,1.08) | 0.20 |  | 0.71 (0.63,0.8) | 2.54E-08 |  | 0.31 | 0.87 (0.78,0.97) | 0.01 |
| rs34136174 | 6 | 32583682 | C | HLA-DQA1 |  | 0.32 | 0.66 (0.58,0.76) | 5.09E-09 |  | 0.35 | 0.86 (0.69,1.08) | 0.20 |  | 0.71 (0.63,0.8) | 2.54E-08 |  | 0.31 | 0.87 (0.78,0.97) | 0.01 |
| rs3129298 | 6 | 32925433 | T | HLA-DMA |  | 0.05 | 0.51 (0.39,0.68) | 1.3E-06 |  | 0.05 | 0.56 (0.37,0.84) | 0.01 |  | 0.53 (0.42,0.66) | 2.79E-08 |  | 0.01 | 0.87 (0.55,1.4) | 0.57 |
| rs28718904 | 6 | 32587687 | G | HLA-DQA1 |  | 0.08 | 0.5 (0.4,0.63) | 3.04E-10 |  | 0.13 | 0.92 (0.68,1.24) | 0.58 |  | 0.62 (0.52,0.74) | 2.92E-08 |  | 0.13 | 0.93 (0.8,1.08) | 0.33 |
| rs34028938 | 6 | 32584346 | A | HLA-DQA1 |  | 0.32 | 0.66 (0.57,0.76) | 3.71E-09 |  | 0.36 | 0.88 (0.7,1.1) | 0.25 |  | 0.72 (0.64,0.81) | 2.97E-08 |  | 0.31 | 0.87 (0.78,0.97) | 0.01 |
| rs35940802 | 6 | 32584355 | G | HLA-DQA1 |  | 0.32 | 0.66 (0.57,0.76) | 3.71E-09 |  | 0.36 | 0.88 (0.7,1.1) | 0.25 |  | 0.72 (0.64,0.81) | 2.97E-08 |  | 0.31 | 0.87 (0.78,0.97) | 0.01 |
| rs35534739 | 6 | 32583490 | G | HLA-DQA1 |  | 0.32 | 0.67 (0.58,0.77) | 9.1E-09 |  | 0.35 | 0.86 (0.68,1.07) | 0.17 |  | 0.71 (0.63,0.8) | 3.08E-08 |  | 0.31 | 0.87 (0.78,0.97) | 0.01 |
| rs34341844 | 6 | 32583496 | A | HLA-DQA1 |  | 0.32 | 0.67 (0.58,0.77) | 9.11E-09 |  | 0.35 | 0.86 (0.68,1.07) | 0.17 |  | 0.71 (0.63,0.8) | 3.08E-08 |  | 0.31 | 0.87 (0.78,0.97) | 0.01 |
| rs15912 | 6 | 32944094 | C | BRD2 (inside gene) |  | 0.05 | 0.51 (0.39,0.68) | 1.1E-06 |  | 0.05 | 0.56 (0.37,0.86) | 0.01 |  | 0.53 (0.42,0.66) | 3.13E-08 |  | 0.01 | 0.87 (0.55,1.4) | 0.57 |
| rs1431394 | 6 | 32923993 | G | HLA-DMA (inside gene) |  | 0.05 | 0.51 (0.39,0.68) | 1.47E-06 |  | 0.05 | 0.56 (0.37,0.84) | 0.01 |  | 0.53 (0.42,0.66) | 3.13E-08 |  | 0.01 | 0.87 (0.55,1.4) | 0.57 |
| rs1431393 | 6 | 32923988 | A | HLA-DMA (inside gene) |  | 0.05 | 0.51 (0.39,0.68) | 1.47E-06 |  | 0.05 | 0.56 (0.37,0.84) | 0.01 |  | 0.53 (0.42,0.66) | 3.13E-08 |  | 0.01 | 0.87 (0.55,1.4) | 0.57 |
| rs3135390 | 6 | 32412395 | C | HLA-DRA (inside gene) |  | 0.27 | 1.63 (1.38,1.91) | 3.71E-09 |  | 0.25 | 1.15 (0.9,1.48) | 0.26 |  | 1.47 (1.28,1.68) | 3.21E-08 |  | 0.27 | 1.36 (1.2,1.54) | 1.52E-06 |
| rs3135034 | 6 | 32951662 | C |  |  | 0.05 | 0.51 (0.39,0.68) | 1.25E-06 |  | 0.05 | 0.56 (0.37,0.86) | 0.01 |  | 0.53 (0.42,0.67) | 3.35E-08 |  | 0.01 | 0.87 (0.55,1.4) | 0.57 |
| rs3097646 | 6 | 32954689 | C |  |  | 0.05 | 0.51 (0.39,0.68) | 1.25E-06 |  | 0.05 | 0.56 (0.37,0.86) | 0.01 |  | 0.53 (0.42,0.67) | 3.35E-08 |  | 0.01 | 0.88 (0.56,1.4) | 0.60 |
| rs4640926 | 6 | 32446993 | T | HLA-DRB9 |  | 0.2 | 1.71 (1.42,2.06) | 1.06E-08 |  | 0.15 | 1.26 (0.9,1.76) | 0.17 |  | 1.59 (1.35,1.87) | 3.49E-08 |  | 0.03 | 1.23 (0.87,1.75) | 0.24 |
| rs34077986 | 6 | 32584191 | C | HLA-DQA1 |  | 0.32 | 0.66 (0.58,0.76) | 5.52E-09 |  | 0.37 | 0.87 (0.7,1.09) | 0.24 |  | 0.72 (0.64,0.81) | 3.69E-08 |  | 0.31 | 0.87 (0.78,0.97) | 0.01 |
| rs3130595 | 6 | 32914297 | A |  |  | 0.05 | 0.52 (0.39,0.68) | 1.75E-06 |  | 0.05 | 0.56 (0.37,0.84) | 0.01 |  | 0.53 (0.42,0.67) | 3.7E-08 |  | 0.01 | 0.87 (0.55,1.4) | 0.57 |
| rs1367727 | 6 | 32934409 | C | HLA-DMA (inside gene) |  | 0.05 | 0.52 (0.39,0.68) | 1.75E-06 |  | 0.05 | 0.56 (0.37,0.84) | 0.01 |  | 0.53 (0.42,0.67) | 3.7E-08 |  | 0.01 | 0.87 (0.55,1.4) | 0.57 |
| rs9269243 | 6 | 32453795 | A | HLA-DRB9 |  | 0.2 | 1.69 (1.4,2.03) | 2.21E-08 |  | 0.15 | 1.29 (0.92,1.81) | 0.13 |  | 1.59 (1.35,1.87) | 3.88E-08 |  | 0.03 | 1.23 (0.87,1.75) | 0.24 |
| rs4959103 | 6 | 32583125 | C | HLA-DQA1 |  | 0.32 | 0.66 (0.58,0.76) | 6.68E-09 |  | 0.36 | 0.87 (0.7,1.09) | 0.23 |  | 0.72 (0.64,0.81) | 4.12E-08 |  | 0.31 | 0.87 (0.78,0.97) | 0.01 |
| rs35542934 | 6 | 32583930 | C | HLA-DQA1 |  | 0.32 | 0.66 (0.58,0.76) | 5.56E-09 |  | 0.37 | 0.88 (0.7,1.1) | 0.25 |  | 0.72 (0.64,0.81) | 4.2E-08 |  | 0.31 | 0.87 (0.78,0.97) | 0.01 |
| rs34850435 | 6 | 32583299 | T | HLA-DQA1 |  | 0.32 | 0.66 (0.58,0.76) | 5.69E-09 |  | 0.37 | 0.88 (0.71,1.1) | 0.26 |  | 0.72 (0.64,0.81) | 4.35E-08 |  | 0.31 | 0.87 (0.78,0.97) | 0.01 |
| rs3135387 | 6 | 32415109 | T |  |  | 0.27 | 1.62 (1.38,1.91) | 4.31E-09 |  | 0.25 | 1.14 (0.89,1.46) | 0.29 |  | 1.46 (1.27,1.67) | 4.37E-08 |  | 0.36 | 1.17 (1.04,1.3) | 0.01 |
| rs4959104 | 6 | 32583129 | C | HLA-DQA1 |  | 0.32 | 0.66 (0.58,0.76) | 6.68E-09 |  | 0.36 | 0.88 (0.7,1.09) | 0.24 |  | 0.72 (0.64,0.81) | 4.41E-08 |  | 0.31 | 0.87 (0.78,0.97) | 0.01 |
| rs35029150 | 6 | 32583426 | G | HLA-DQA1 |  | 0.32 | 0.66 (0.58,0.76) | 5.84E-09 |  | 0.37 | 0.88 (0.71,1.1) | 0.26 |  | 0.72 (0.64,0.81) | 4.42E-08 |  | 0.31 | 0.87 (0.78,0.97) | 0.01 |
| rs35128369 | 6 | 32583403 | G | HLA-DQA1 |  | 0.32 | 0.66 (0.58,0.76) | 5.84E-09 |  | 0.37 | 0.88 (0.71,1.1) | 0.26 |  | 0.72 (0.64,0.81) | 4.42E-08 |  | 0.31 | 0.87 (0.78,0.97) | 0.02 |
| rs34985232 | 6 | 32584015 | G | HLA-DQA1 |  | 0.32 | 0.66 (0.58,0.76) | 5.6E-09 |  | 0.37 | 0.88 (0.71,1.1) | 0.26 |  | 0.72 (0.64,0.81) | 4.49E-08 |  | 0.31 | 0.87 (0.78,0.97) | 0.01 |
| rs35928237 | 6 | 32583461 | T | HLA-DQA1 |  | 0.32 | 0.66 (0.58,0.76) | 5.68E-09 |  | 0.37 | 0.88 (0.71,1.1) | 0.26 |  | 0.72 (0.64,0.81) | 4.53E-08 |  | 0.31 | 0.87 (0.77,0.97) | 0.01 |
| rs4959106 | 6 | 32583159 | C | HLA-DQA1 |  | 0.32 | 0.66 (0.58,0.76) | 5.69E-09 |  | 0.37 | 0.88 (0.71,1.1) | 0.26 |  | 0.72 (0.64,0.81) | 4.54E-08 |  | 0.31 | 0.87 (0.78,0.97) | 0.01 |
| rs75518178 | 6 | 32505100 | T | HLA-DRB5 |  | 0.35 | 0.62 (0.52,0.73) | 1.18E-08 |  | 0.35 | 0.83 (0.63,1.1) | 0.20 |  | 0.67 (0.58,0.77) | 4.75E-08 |  | 0.22 | 0.85 (0.62,1.15) | 0.29 |
| rs34537691 | 6 | 32584295 | G | HLA-DQA1 |  | 0.32 | 0.66 (0.58,0.76) | 6.2E-09 |  | 0.37 | 0.88 (0.71,1.1) | 0.26 |  | 0.72 (0.64,0.81) | 4.87E-08 |  | 0.31 | 0.87 (0.78,0.97) | 0.01 |
| SNP: single nucleotide polymorphism; Chr: chromosome; MAF: minor allele frequency; OR: odds ratio; 95% CI: 95% confidence interval  *The result of rs13204736 in the African American Cohort did not pass the imputation quality control. The result was obtained from the sequencing data on 932 subjects.  **The minor allele for rs2395206 in African American Cohort is T and the odds ratio is calculated based on allele T | | | | | | | | | | | | | | | | | | | |

| **Table S2**: Odds ratios and p-values for the top 7 SNP adjusted for other significant SNPs using stepwise approach | | | | | | | | | | | | |
| --- | --- | --- | --- | --- | --- | --- | --- | --- | --- | --- | --- | --- |
|  | | | | | Phase 1 | | | Phase 2 | | | Meta-analysis | |
| SNP | Chr | Position | Minor allele | Nearest gene (+/-25K) | MAF case* | OR (95% CI) | p-value | MAF case | OR (95% CI) | p-value | OR (95% CI) | p-value |
| Step 1: adjusted for the most significant SNPs in the original model, rs9269233 | | | | | | | | | | | | |
| rs9276935 | 6 | 32936441 | T | BRD2 | 0.05 | 0.53 (0.40, 0.70) | 9.47E-06 | 0.05 | 0.56 (0.37, 0.84) | 5.66E-03 | 0.54 (0.42, 0.68) | 1.79E-07 |
| rs28589559 | 6 | 32587716 | G | HLA-DQA1 | 0.08 | 0.59 (0.47, 0.75) | 7.53E-06 | 0.13 | 0.93 (0.68, 1.28) | 0.66 | 0.69 (0.58, 0.83) | 7.08E-05 |
| rs71549283 | 6 | 32505038 | G | HLA-DRB5 | 0.33 | 0.74 (0.62, 0.90) | 2.01E-03 | 0.34 | 0.84 (0.63, 1.13) | 0.26 | 0.77 (0.66, 0.90) | 1.38E-03 |
| rs35656642 | 6 | 32583610 | C | HLA-DQA1 | 0.32 | 0.80 (0.68, 0.93) | 4.15E-03 | 0.34 | 0.84 (0.66, 1.07) | 0.16 | 0.81 (0.71, 0.92) | 1.53E-03 |
| rs9271346 | 6 | 32583468 | C | HLA-DQA1 | 0.27 | 1.36 (1.12, 1.65) | 1.91E-03 | 0.24 | 1.13 (0.85, 1.50) | 0.40 | 1.28 (1.09, 1.50) | 2.33E-03 |
| rs3129888 | 6 | 32411726 | G | HLA-DRA | 0.27 | 1.22 (0.99, 1.50) | 0.06 | 0.25 | 1.15 (0.86, 1.55) | 0.35 | 1.20 (1.01, 1.42) | 0.04 |
| Step 2: adjusted for the most significant SNPs in the original model and step 1, rs9269233 and rs9276935 | | | | | | | | | | | | |
| rs28589559 | 6 | 32587716 | G | HLA-DQA1 | 0.08 | 0.59 (0.47, 0.74) | 5.41E-06 | 0.13 | 0.93 (0.68, 1.27) | 0.64 | 0.69 (0.57, 0.83) | 5.35E-05 |
| rs71549283 | 6 | 32505038 | G | HLA-DRB5 | 0.33 | 0.77 (0.64, 0.93) | 7.30E-03 | 0.34 | 0.85 (0.63, 1.15) | 0.29 | 0.79 (0.68, 0.93) | 4.83E-03 |
| rs9271346 | 6 | 32583468 | C | HLA-DQA1 | 0.27 | 1.32 (1.08, 1.60) | 5.54E-03 | 0.24 | 1.14 (0.85, 1.51) | 0.38 | 1.26 (1.07, 1.48) | 5.16E-03 |
| rs35656642 | 6 | 32583610 | C | HLA-DQA1 | 0.32 | 0.83 (0.71, 0.97) | 0.02 | 0.34 | 0.87 (0.68, 1.11) | 0.26 | 0.84 (0.73, 0.96) | 0.01 |
| rs3129888 | 6 | 32411726 | G | HLA-DRA | 0.27 | 1.19 (0.96, 1.46) | 0.11 | 0.25 | 1.16 (0.86, 1.55) | 0.34 | 1.18 (0.99, 1.39) | 0.06 |
| Step 3: adjusted for the most significant SNPs in the original model, step 1, and step 2, rs9269233, rs9276935, and rs28589559 | | | | | | | | | | | | |
| rs3129888 | 6 | 32411726 | G | HLA-DRA | 0.27 | 1.30 (1.05, 1.60) | 0.02 | 0.25 | 1.17 (0.87, 1.58) | 0.30 | 1.25 (1.06, 1.49) | 9.58E-03 |
| rs9271346 | 6 | 32583468 | C | HLA-DQA1 | 0.27 | 1.28 (1.05, 1.55) | 0.02 | 0.24 | 1.13 (0.85, 1.51) | 0.40 | 1.23 (1.04, 1.44) | 0.01 |
| rs71549283 | 6 | 32505038 | G | HLA-DRB5 | 0.33 | 0.81 (0.67, 0.97) | 0.03 | 0.34 | 0.86 (0.63, 1.17) | 0.33 | 0.82 (0.70, 0.96) | 0.02 |
| rs35656642 | 6 | 32583610 | C | HLA-DQA1 | 0.32 | 0.96 (0.81, 1.15) | 0.67 | 0.34 | 0.87 (0.67, 1.13) | 0.31 | 0.93 (0.81, 1.08) | 0.35 |
| SNP: single nucleotide polymorphism; Chr: chromosome; MAF: minor allele frequency; OR: odds ratio; 95% CI: 95% confidence interval | | | | | | | | | | | | |

| **Table S3:** Odds ratios and p-values of other SNPs with p-value <5 x 10^-5^ (but >5x10^-8^). | | | | | | | | | | | | |
| --- | --- | --- | --- | --- | --- | --- | --- | --- | --- | --- | --- | --- |
|  |  |  |  |  | Phase1 | | | Phase 2 | | | Meta-analysis | |
| SNP | Chr | Position | Minor allele | Nearest gene | MAF case | OR (95% CI) | p-value | MAF case | OR (95% CI) | p-value | OR (95% CI) | p-value |
| rs2395202 | 6 | 32451906 | T | HLA-DRB9 | 0.44 | 1.68 (1.46,1.94) | 5.99e-13 | 0.4 | 1.03 (0.82,1.28) | 8.20e-01 | 1.45 (1.29,1.64) | 9.54e-10 |
| 6 | 6 | 32452802 | T | HLA-DRB9 | 0.42 | 1.65 (1.42,1.93) | 5.18e-11 | 0.39 | 1.06 (0.84,1.35) | 6.02e-01 | 1.45 (1.28,1.65) | 8.82e-09 |
| rs28359884 | 6 | 32584330 | A | HLA-DQA1 | 0.43 | 0.65 (0.57,0.75) | 2.30e-09 | 0.47 | 0.91 (0.73,1.14) | 4.20e-01 | 0.72 (0.64,0.81) | 6.00e-08 |
| rs114360309 | 6 | 32626377 | A | CMAHP | 0.04 | 0.38 (0.29,0.52) | 1.33e-11 | 0.07 | 1.09 (0.72,1.64) | 6.89e-01 | 0.55 (0.43,0.69) | 6.44e-08 |
| rs200017582 | 6 | 32568090 | C | HLA-DRB1 | 0.07 | 0.45 (0.36,0.58) | 2.51e-11 | 0.11 | 1.05 (0.74,1.49) | 7.90e-01 | 0.6 (0.49,0.73) | 6.53e-08 |
| 6 | 6 | 32569459 | A | HLA-DRB1 | 0.07 | 0.46 (0.36,0.59) | 4.55e-11 | 0.11 | 1.03 (0.73,1.43) | 8.85e-01 | 0.6 (0.5,0.73) | 6.74e-08 |
| rs3129752 | 6 | 32582627 | C | HLA-DQA1 | 0.32 | 0.67 (0.58,0.77) | 1.14e-08 | 0.36 | 0.88 (0.7,1.09) | 2.44e-01 | 0.72 (0.64,0.81) | 6.83e-08 |
| rs9270596 | 6 | 32561582 | T | HLA-DRB1 | 0.21 | 1.66 (1.38,2) | 4.95e-08 | 0.16 | 1.29 (0.93,1.8) | 1.25e-01 | 1.57 (1.33,1.84) | 7.09e-08 |
| rs112627226 | 6 | 32582969 | C | HLA-DQA1 | 0.04 | 0.38 (0.29,0.51) | 9.64e-12 | 0.07 | 1.11 (0.74,1.68) | 6.15e-01 | 0.55 (0.43,0.7) | 7.10e-08 |
| rs9268925 | 6 | 32432969 | A | HLA-DRB9 | 0.2 | 1.71 (1.42,2.06) | 1.06e-08 | 0.17 | 1.19 (0.88,1.61) | 2.57e-01 | 1.55 (1.32,1.81) | 7.11e-08 |
| rs9270187 | 6 | 32555965 | T | HLA-DRB1 | 0.2 | 1.71 (1.41,2.07) | 2.33e-08 | 0.15 | 1.26 (0.89,1.78) | 1.82e-01 | 1.59 (1.35,1.88) | 7.17e-08 |
| rs9270193 | 6 | 32556157 | A | HLA-DRB1 | 0.2 | 1.7 (1.41,2.06) | 2.43e-08 | 0.16 | 1.26 (0.9,1.77) | 1.79e-01 | 1.59 (1.35,1.87) | 7.18e-08 |
| 6 | 6 | 32473455 | T | HLA-DRB5 | 0.04 | 0.36 (0.26,0.5) | 9.98e-12 | 0.07 | 1.11 (0.74,1.66) | 6.18e-01 | 0.55 (0.43,0.71) | 7.19e-08 |
| rs67505275 | 6 | 32449386 | A | HLA-DRB9 | 0.04 | 0.4 (0.3,0.53) | 3.14e-11 | 0.07 | 1.06 (0.69,1.62) | 7.98e-01 | 0.54 (0.43,0.69) | 7.37e-08 |
| rs57425824 | 6 | 32586011 | T | HLA-DQA1 | 0.04 | 0.38 (0.29,0.51) | 1.04e-11 | 0.07 | 1.11 (0.74,1.68) | 6.15e-01 | 0.55 (0.43,0.7) | 7.48e-08 |
| rs78660114 | 6 | 32452767 | G | HLA-DRB9 | 0.54 | 1.59 (1.38,1.83) | 7.22e-11 | 0.48 | 1.01 (0.81,1.25) | 9.38e-01 | 1.38 (1.23,1.56) | 7.53e-08 |
| rs28535317 | 6 | 32589065 | G | HLA-DQA1 | 0.04 | 0.38 (0.29,0.52) | 1.09e-11 | 0.07 | 1.11 (0.74,1.68) | 6.15e-01 | 0.55 (0.43,0.7) | 7.73e-08 |
| rs9269160 | 6 | 32446711 | T | HLA-DRB9 | 0.53 | 1.63 (1.41,1.88) | 1.44e-11 | 0.48 | 0.95 (0.77,1.18) | 6.50e-01 | 1.38 (1.23,1.56) | 8.00e-08 |
| rs28752515 | 6 | 32583603 | G | HLA-DQA1 | 0.54 | 1.52 (1.32,1.74) | 2.87e-09 | 0.49 | 0.92 (0.74,1.14) | 4.47e-01 | 1.32 (1.17,1.48) | 8.20e-08 |
| rs35168843 | 6 | 32442408 | A | HLA-DRB9 | 0.04 | 0.38 (0.28,0.51) | 9.81e-12 | 0.07 | 1.12 (0.75,1.68) | 5.86e-01 | 0.56 (0.44,0.71) | 8.20e-08 |
| 6 | 6 | 32452346 | C | HLA-DRB9 | 0.05 | 0.4 (0.3,0.52) | 5.06e-12 | 0.08 | 1.15 (0.77,1.72) | 4.92e-01 | 0.56 (0.45,0.71) | 8.22e-08 |
| rs9271065 | 6 | 32575619 | C | HLA-DRB1 | 0.2 | 1.68 (1.4,2.03) | 2.49e-08 | 0.17 | 1.22 (0.9,1.64) | 1.93e-01 | 1.54 (1.31,1.8) | 8.37e-08 |
| rs147716103 | 6 | 32445319 | C | HLA-DRB9 | 0.04 | 0.39 (0.29,0.52) | 1.46e-11 | 0.07 | 1.1 (0.73,1.66) | 6.36e-01 | 0.55 (0.44,0.7) | 8.57e-08 |
| rs28752514 | 6 | 32583576 | T | HLA-DQA1 | 0.54 | 1.51 (1.32,1.74) | 3.44e-09 | 0.49 | 0.92 (0.73,1.14) | 4.35e-01 | 1.31 (1.17,1.48) | 8.84e-08 |
| rs28864514 | 6 | 32569431 | G | HLA-DRB1 | 0.07 | 0.46 (0.36,0.59) | 4.60e-11 | 0.11 | 1.04 (0.74,1.45) | 8.17e-01 | 0.61 (0.5,0.74) | 8.87e-08 |
| rs34442410 | 6 | 32442086 | G | HLA-DRB9 | 0.04 | 0.37 (0.27,0.5) | 6.57e-12 | 0.07 | 1.15 (0.76,1.75) | 4.93e-01 | 0.55 (0.43,0.71) | 9.66e-08 |
| rs3101944 | 6 | 32911127 | T | XXbac-BPG181M17.5 | 0.05 | 0.53 (0.41,0.7) | 3.73e-06 | 0.05 | 0.57 (0.38,0.86) | 7.65e-03 | 0.54 (0.43,0.68) | 9.92e-08 |
| rs28833271 | 6 | 32585045 | T | HLA-DQA1 | 0.05 | 0.46 (0.35,0.6) | 1.29e-09 | 0.08 | 1.02 (0.7,1.48) | 9.16e-01 | 0.6 (0.48,0.74) | 6.02e-07 |
| rs9269037 | 6 | 32437958 | T | HLA-DRB9 | 0.43 | 1.58 (1.37,1.82) | 3.12e-10 | 0.37 | 0.95 (0.75,1.19) | 6.58e-01 | 1.37 (1.21,1.54) | 6.03e-07 |
| rs9272460 | 6 | 32605583 | G | HLA-DQA1 | 0.24 | 0.63 (0.54,0.73) | 1.70e-09 | 0.28 | 1 (0.79,1.28) | 9.68e-01 | 0.72 (0.63,0.82) | 6.06e-07 |
| rs34053446 | 6 | 32586809 | G | HLA-DQA1 | 0.05 | 0.46 (0.35,0.6) | 1.27e-09 | 0.08 | 1.02 (0.71,1.48) | 9.08e-01 | 0.6 (0.48,0.74) | 6.15e-07 |
| rs67294117 | 6 | 32504774 | A | HLA-DRB5 | 0.05 | 0.42 (0.31,0.58) | 5.92e-09 | 0.09 | 0.95 (0.65,1.39) | 7.95e-01 | 0.58 (0.46,0.74) | 6.18e-07 |
| rs3117098 | 6 | 32358513 | G | HCG23 | 0.4 | 1.52 (1.32,1.75) | 7.59e-09 | 0.37 | 1.04 (0.83,1.3) | 7.47e-01 | 1.36 (1.2,1.53) | 6.18e-07 |
| rs3129952 | 6 | 32359763 | G | HCG23 | 0.4 | 1.52 (1.32,1.75) | 7.60e-09 | 0.37 | 1.04 (0.83,1.3) | 7.47e-01 | 1.36 (1.2,1.53) | 6.18e-07 |
| rs35826015 | 6 | 32590669 | A | HLA-DQA1 | 0.05 | 0.46 (0.35,0.6) | 1.43e-09 | 0.08 | 1.02 (0.7,1.47) | 9.28e-01 | 0.6 (0.48,0.74) | 6.20e-07 |
| rs13202749 | 6 | 32585299 | A | HLA-DQA1 | 0.05 | 0.46 (0.35,0.6) | 1.32e-09 | 0.08 | 1.02 (0.71,1.48) | 9.11e-01 | 0.6 (0.48,0.74) | 6.23e-07 |
| rs17533167 | 6 | 32590844 | C | HLA-DQA1 | 0.05 | 0.46 (0.35,0.6) | 1.45e-09 | 0.08 | 1.02 (0.7,1.47) | 9.28e-01 | 0.6 (0.48,0.74) | 6.24e-07 |
| rs3129898 | 6 | 32422125 | G | HLA-DRB9 | 0.2 | 1.66 (1.39,2) | 3.84e-08 | 0.17 | 1.12 (0.83,1.5) | 4.61e-01 | 1.49 (1.27,1.74) | 6.25e-07 |
| rs17599077 | 6 | 32591058 | C | HLA-DQA1 | 0.05 | 0.46 (0.35,0.6) | 1.46e-09 | 0.08 | 1.02 (0.7,1.47) | 9.29e-01 | 0.6 (0.48,0.74) | 6.25e-07 |
| rs72847586 | 6 | 32570172 | C | HLA-DRB1 | 0.05 | 0.46 (0.35,0.6) | 1.45e-09 | 0.08 | 1.02 (0.7,1.47) | 9.28e-01 | 0.6 (0.48,0.74) | 6.26e-07 |
| rs35150023 | 6 | 32586754 | A | HLA-DQA1 | 0.05 | 0.46 (0.35,0.6) | 1.33e-09 | 0.08 | 1.02 (0.71,1.48) | 9.08e-01 | 0.6 (0.48,0.74) | 6.34e-07 |
| rs28872589 | 6 | 32584989 | C | HLA-DQA1 | 0.05 | 0.46 (0.35,0.6) | 1.32e-09 | 0.08 | 1.02 (0.71,1.48) | 9.06e-01 | 0.6 (0.48,0.74) | 6.34e-07 |
| rs9270140 | 6 | 32555206 | A | HLA-DRB1 | 0.08 | 0.53 (0.42,0.66) | 1.13e-08 | 0.12 | 0.93 (0.67,1.3) | 6.81e-01 | 0.63 (0.52,0.76) | 6.41e-07 |
| rs9270139 | 6 | 32555203 | C | HLA-DRB1 | 0.08 | 0.53 (0.42,0.66) | 1.13e-08 | 0.12 | 0.93 (0.67,1.3) | 6.81e-01 | 0.63 (0.52,0.76) | 6.41e-07 |
| rs9268844 | 6 | 32429153 | AT | HLA-DRB9 | 0.2 | 0.66 (0.57,0.78) | 2.78e-07 | 0.21 | 0.85 (0.66,1.09) | 2.05e-01 | 0.71 (0.62,0.81) | 6.46e-07 |
| rs3117108 | 6 | 32342822 | C | CMAHP | 0.4 | 1.52 (1.31,1.75) | 9.34e-09 | 0.37 | 1.04 (0.83,1.3) | 7.20e-01 | 1.36 (1.2,1.53) | 6.48e-07 |
| rs113500036 | 6 | 32584060 | G | HLA-DQA1 | 0.05 | 0.46 (0.36,0.6) | 1.55e-09 | 0.08 | 1.02 (0.7,1.47) | 9.28e-01 | 0.6 (0.49,0.74) | 6.54e-07 |
| rs72847591 | 6 | 32570494 | T | HLA-DRB1 | 0.06 | 0.46 (0.36,0.6) | 1.51e-09 | 0.08 | 1.02 (0.7,1.47) | 9.23e-01 | 0.6 (0.49,0.74) | 6.55e-07 |
| rs77632534 | 6 | 32576283 | A | HLA-DRB1 | 0.05 | 0.46 (0.35,0.6) | 1.57e-09 | 0.08 | 1.02 (0.7,1.47) | 9.30e-01 | 0.6 (0.48,0.74) | 6.56e-07 |
| rs34811813 | 6 | 32576265 | T | HLA-DRB1 | 0.05 | 0.46 (0.35,0.6) | 1.57e-09 | 0.08 | 1.02 (0.7,1.47) | 9.30e-01 | 0.6 (0.48,0.74) | 6.57e-07 |
| rs28383268 | 6 | 32585982 | T | HLA-DQA1 | 0.05 | 0.46 (0.35,0.6) | 1.33e-09 | 0.08 | 1.03 (0.71,1.49) | 8.93e-01 | 0.6 (0.49,0.74) | 6.69e-07 |
| rs3763323 | 6 | 32406843 | C | HLA-DRA | 0.21 | 1.63 (1.36,1.95) | 1.39e-07 | 0.17 | 1.17 (0.87,1.56) | 2.90e-01 | 1.48 (1.27,1.73) | 6.71e-07 |
| rs36006606 | 6 | 32591424 | G | HLA-DQA1 | 0.05 | 0.45 (0.34,0.59) | 1.86e-09 | 0.08 | 1.01 (0.7,1.46) | 9.56e-01 | 0.6 (0.48,0.75) | 6.73e-07 |
| rs4999342 | 6 | 32448098 | T | HLA-DRB9 | 0.39 | 1.57 (1.35,1.81) | 1.24e-09 | 0.35 | 0.98 (0.78,1.24) | 8.74e-01 | 1.37 (1.21,1.55) | 6.82e-07 |
| rs3117104 | 6 | 32348309 | T | CMAHP | 0.4 | 1.52 (1.31,1.75) | 9.50e-09 | 0.37 | 1.04 (0.83,1.3) | 7.31e-01 | 1.36 (1.2,1.53) | 6.85e-07 |
| rs36206696 | 6 | 32561009 | C | HLA-DRB1 | 0.05 | 0.45 (0.35,0.59) | 9.66e-10 | 0.08 | 1.04 (0.71,1.53) | 8.25e-01 | 0.59 (0.48,0.74) | 6.87e-07 |
| rs3129957 | 6 | 32368593 | A | BTNL2 | 0.4 | 1.51 (1.31,1.75) | 1.04e-08 | 0.37 | 1.04 (0.83,1.3) | 7.15e-01 | 1.36 (1.2,1.53) | 6.89e-07 |
| rs6931044 | 6 | 32583194 | T | HLA-DQA1 | 0.31 | 0.69 (0.6,0.79) | 1.66e-07 | 0.34 | 0.88 (0.71,1.1) | 2.72e-01 | 0.74 (0.66,0.83) | 6.91e-07 |
| rs3117102 | 6 | 32350107 | C | HCG23 | 0.4 | 1.52 (1.31,1.75) | 9.53e-09 | 0.37 | 1.04 (0.83,1.3) | 7.33e-01 | 1.36 (1.2,1.53) | 6.92e-07 |
| rs3117105 | 6 | 32343714 | T | CMAHP | 0.4 | 1.51 (1.31,1.75) | 1.04e-08 | 0.37 | 1.04 (0.83,1.3) | 7.18e-01 | 1.36 (1.2,1.53) | 6.94e-07 |
| rs3129946 | 6 | 32346772 | A | CMAHP | 0.4 | 1.52 (1.31,1.75) | 9.53e-09 | 0.37 | 1.04 (0.83,1.3) | 7.35e-01 | 1.36 (1.2,1.53) | 6.95e-07 |
| rs34113780 | 6 | 32575258 | G | HLA-DRB1 | 0.05 | 0.46 (0.35,0.6) | 1.32e-09 | 0.08 | 1.03 (0.71,1.49) | 8.81e-01 | 0.6 (0.49,0.74) | 6.95e-07 |
| rs73405151 | 6 | 32486756 | T | HLA-DRB5 | 0.21 | 1.61 (1.34,1.93) | 2.37e-07 | 0.18 | 1.19 (0.89,1.6) | 2.32e-01 | 1.48 (1.27,1.73) | 6.97e-07 |
| rs9268468 | 6 | 32353166 | A | HCG23 | 0.4 | 1.52 (1.31,1.75) | 9.94e-09 | 0.37 | 1.04 (0.83,1.3) | 7.29e-01 | 1.36 (1.2,1.53) | 7.02e-07 |
| rs73575927 | 19 | 52985887 | C | ZNF578 | 0.13 | 1.89 (1.43,2.5) | 4.66e-06 | 0.11 | 1.69 (1,2.83) | 3.83e-02 | 1.84 (1.44,2.36) | 7.07e-07 |
| rs7749092 | 6 | 32449050 | T | HLA-DRB9 | 0.39 | 1.58 (1.36,1.83) | 1.12e-09 | 0.36 | 0.98 (0.78,1.23) | 8.40e-01 | 1.37 (1.21,1.55) | 7.22e-07 |
| rs36233210 | 6 | 32577417 | G | HLA-DQA1 | 0.05 | 0.46 (0.35,0.6) | 1.31e-09 | 0.08 | 1.03 (0.71,1.49) | 8.66e-01 | 0.6 (0.49,0.75) | 7.31e-07 |
| rs189570112 | 6 | 32565711 | C | HLA-DRB1 | 0.05 | 0.45 (0.35,0.59) | 9.48e-10 | 0.08 | 1.05 (0.72,1.54) | 8.02e-01 | 0.59 (0.48,0.74) | 7.38e-07 |
| rs34350660 | 6 | 32550491 | T | HLA-DRB1 | 0.05 | 0.46 (0.35,0.59) | 1.34e-09 | 0.07 | 1.04 (0.69,1.55) | 8.67e-01 | 0.58 (0.47,0.73) | 7.40e-07 |
| rs72850250 | 6 | 32550252 | A | HLA-DRB1 | 0.06 | 0.46 (0.35,0.6) | 1.72e-09 | 0.08 | 1.02 (0.69,1.51) | 9.13e-01 | 0.59 (0.48,0.74) | 7.43e-07 |
| rs111665670 | 6 | 32567065 | A | HLA-DRB1 | 0.05 | 0.45 (0.35,0.59) | 1.00e-09 | 0.08 | 1.05 (0.71,1.54) | 8.07e-01 | 0.59 (0.48,0.74) | 7.52e-07 |
| rs9268464 | 6 | 32352568 | T | HCG23 | 0.4 | 1.51 (1.31,1.75) | 1.09e-08 | 0.37 | 1.04 (0.83,1.3) | 7.31e-01 | 1.36 (1.2,1.53) | 7.53e-07 |
| rs35259208 | 6 | 32550548 | G | HLA-DRB1 | 0.05 | 0.46 (0.35,0.59) | 1.38e-09 | 0.07 | 1.04 (0.69,1.55) | 8.67e-01 | 0.58 (0.47,0.73) | 7.55e-07 |
| rs35662241 | 6 | 32550580 | T | HLA-DRB1 | 0.05 | 0.46 (0.35,0.59) | 1.38e-09 | 0.07 | 1.04 (0.69,1.55) | 8.67e-01 | 0.58 (0.47,0.73) | 7.55e-07 |
| rs34981130 | 6 | 32546670 | T | HLA-DRB1 | 0.06 | 0.45 (0.35,0.59) | 9.75e-10 | 0.09 | 1.05 (0.73,1.51) | 7.99e-01 | 0.6 (0.49,0.75) | 7.61e-07 |
| rs17205002 | 6 | 32578346 | G | HLA-DQA1 | 0.05 | 0.46 (0.35,0.6) | 1.42e-09 | 0.08 | 1.03 (0.71,1.49) | 8.66e-01 | 0.6 (0.49,0.75) | 7.73e-07 |
| rs17211356 | 6 | 32578426 | G | HLA-DQA1 | 0.05 | 0.46 (0.35,0.6) | 1.42e-09 | 0.08 | 1.03 (0.71,1.49) | 8.66e-01 | 0.6 (0.49,0.75) | 7.73e-07 |
| rs35129460 | 6 | 32550785 | C | HLA-DRB1 | 0.05 | 0.45 (0.35,0.59) | 1.37e-09 | 0.08 | 1.04 (0.69,1.55) | 8.57e-01 | 0.58 (0.47,0.73) | 7.75e-07 |
| rs34553538 | 6 | 32564856 | C | HLA-DRB1 | 0.05 | 0.45 (0.34,0.59) | 1.14e-09 | 0.08 | 1.05 (0.71,1.54) | 8.20e-01 | 0.59 (0.48,0.74) | 7.84e-07 |
| rs9272478 | 6 | 32605767 | A | HLA-DQA1 | 0.05 | 0.46 (0.36,0.6) | 2.71e-09 | 0.08 | 1 (0.69,1.45) | 9.84e-01 | 0.6 (0.49,0.75) | 7.91e-07 |
| rs28895204 | 6 | 32425196 | G | HLA-DRB9 | 0.08 | 0.49 (0.39,0.62) | 9.86e-10 | 0.1 | 1.05 (0.74,1.49) | 7.90e-01 | 0.62 (0.51,0.76) | 7.94e-07 |
| rs28895202 | 6 | 32425190 | C | HLA-DRB9 | 0.08 | 0.49 (0.39,0.62) | 9.86e-10 | 0.1 | 1.05 (0.74,1.49) | 7.90e-01 | 0.62 (0.51,0.76) | 7.94e-07 |
| rs28895203 | 6 | 32425191 | A | HLA-DRB9 | 0.08 | 0.49 (0.39,0.62) | 9.86e-10 | 0.1 | 1.05 (0.74,1.49) | 7.90e-01 | 0.62 (0.51,0.76) | 7.94e-07 |
| rs13194888 | 6 | 32425185 | T | HLA-DRB9 | 0.08 | 0.49 (0.39,0.62) | 9.86e-10 | 0.1 | 1.05 (0.74,1.49) | 7.90e-01 | 0.62 (0.51,0.76) | 7.94e-07 |
| rs199903584 | 6 | 32425167 | G | HLA-DRB9 | 0.08 | 0.49 (0.39,0.62) | 9.87e-10 | 0.1 | 1.05 (0.74,1.49) | 7.89e-01 | 0.62 (0.51,0.76) | 7.96e-07 |
| rs34310302 | 6 | 32606597 | A | HLA-DQA1 | 0.05 | 0.46 (0.36,0.6) | 2.44e-09 | 0.08 | 1.01 (0.7,1.46) | 9.60e-01 | 0.6 (0.49,0.75) | 8.00e-07 |
| rs36218421 | 6 | 32550227 | A | HLA-DRB1 | 0.06 | 0.46 (0.35,0.6) | 1.75e-09 | 0.08 | 1.03 (0.7,1.51) | 8.95e-01 | 0.59 (0.48,0.74) | 8.03e-07 |
| rs72847588 | 6 | 32570216 | A | HLA-DRB1 | 0.05 | 0.46 (0.35,0.6) | 1.47e-09 | 0.08 | 1.03 (0.71,1.5) | 8.57e-01 | 0.6 (0.49,0.75) | 8.13e-07 |
| rs72850273 | 6 | 32550816 | A | HLA-DRB1 | 0.05 | 0.46 (0.35,0.6) | 1.60e-09 | 0.08 | 1.03 (0.69,1.55) | 8.69e-01 | 0.59 (0.47,0.73) | 8.29e-07 |
| rs4382332 | 6 | 32572155 | C | HLA-DRB1 | 0.05 | 0.46 (0.36,0.6) | 1.61e-09 | 0.08 | 1.03 (0.71,1.49) | 8.68e-01 | 0.6 (0.49,0.75) | 8.33e-07 |
| rs3129954 | 6 | 32365580 | A | BTNL2 | 0.4 | 1.52 (1.31,1.75) | 9.53e-09 | 0.37 | 1.03 (0.83,1.29) | 7.84e-01 | 1.35 (1.2,1.53) | 8.37e-07 |
| rs72844283 | 6 | 32565189 | T | HLA-DRB1 | 0.05 | 0.45 (0.35,0.59) | 9.49e-10 | 0.08 | 1.06 (0.72,1.55) | 7.68e-01 | 0.6 (0.48,0.74) | 8.39e-07 |
| rs375399031 | 20 | 20336122 | G | C20orf26 | 0.39 | 1.46 (1.23,1.73) | 8.43e-06 | 0.4 | 1.36 (1.03,1.8) | 2.79e-02 | 1.43 (1.24,1.65) | 8.43e-07 |
| rs55783839 | 20 | 20336121 | G | C20orf26 | 0.39 | 1.46 (1.23,1.73) | 8.42e-06 | 0.4 | 1.36 (1.03,1.8) | 2.80e-02 | 1.43 (1.24,1.65) | 8.43e-07 |
| rs202127825 | 6 | 32550406 | C | HLA-DRB1 | 0.05 | 0.46 (0.35,0.6) | 1.64e-09 | 0.07 | 1.03 (0.69,1.55) | 8.68e-01 | 0.59 (0.47,0.73) | 8.45e-07 |
| rs199535050 | 6 | 32550417 | A | HLA-DRB1 | 0.05 | 0.46 (0.35,0.6) | 1.64e-09 | 0.07 | 1.03 (0.69,1.55) | 8.68e-01 | 0.59 (0.47,0.73) | 8.45e-07 |
| rs72847996 | 6 | 32384892 | A | CMAHP | 0.04 | 0.42 (0.32,0.56) | 2.52e-10 | 0.08 | 1.13 (0.76,1.69) | 5.40e-01 | 0.59 (0.47,0.74) | 8.55e-07 |
| rs72847691 | 6 | 32546940 | T | HLA-DRB1 | 0.06 | 0.46 (0.35,0.59) | 1.38e-09 | 0.1 | 1.04 (0.73,1.48) | 8.32e-01 | 0.61 (0.5,0.76) | 8.55e-07 |
| rs112464499 | 6 | 32559467 | G | HLA-DRB1 | 0.05 | 0.44 (0.34,0.58) | 5.85e-10 | 0.09 | 1.09 (0.74,1.59) | 6.68e-01 | 0.6 (0.48,0.74) | 8.84e-07 |
| rs113147629 | 6 | 32566947 | T | HLA-DRB1 | 0.05 | 0.46 (0.35,0.59) | 1.26e-09 | 0.08 | 1.05 (0.72,1.54) | 8.05e-01 | 0.6 (0.48,0.74) | 8.87e-07 |
| rs3129860 | 6 | 32401079 | A | HLA-DRA | 0.2 | 1.62 (1.35,1.94) | 2.00e-07 | 0.17 | 1.17 (0.87,1.56) | 2.90e-01 | 1.47 (1.26,1.72) | 8.97e-07 |
| rs3129865 | 6 | 32404048 | G | HLA-DRA | 0.2 | 1.62 (1.35,1.94) | 2.00e-07 | 0.17 | 1.17 (0.87,1.56) | 2.90e-01 | 1.47 (1.26,1.72) | 8.97e-07 |
| rs3129868 | 6 | 32404377 | A | HLA-DRA | 0.2 | 1.62 (1.35,1.94) | 2.00e-07 | 0.17 | 1.17 (0.87,1.56) | 2.90e-01 | 1.47 (1.26,1.72) | 8.97e-07 |
| rs112604035 | 6 | 32559643 | A | HLA-DRB1 | 0.06 | 0.47 (0.36,0.61) | 3.42e-09 | 0.09 | 1 (0.69,1.46) | 9.89e-01 | 0.6 (0.48,0.74) | 9.15e-07 |
| rs199927008 | 6 | 32549627 | C | HLA-DRB1 | 0.05 | 0.46 (0.35,0.6) | 1.87e-09 | 0.07 | 1.03 (0.69,1.55) | 8.69e-01 | 0.59 (0.47,0.73) | 9.21e-07 |
| rs72847660 | 6 | 32544308 | T | HLA-DRB1 | 0.05 | 0.45 (0.34,0.59) | 1.67e-09 | 0.09 | 1.04 (0.72,1.5) | 8.44e-01 | 0.6 (0.49,0.75) | 9.34e-07 |
| rs17211657 | 6 | 32603023 | A | HLA-DQA1 | 0.05 | 0.46 (0.36,0.6) | 2.67e-09 | 0.08 | 1.02 (0.7,1.47) | 9.34e-01 | 0.6 (0.49,0.75) | 9.35e-07 |
| rs3817967 | 6 | 32361469 | T | HCG23 | 0.08 | 0.49 (0.39,0.62) | 1.20e-10 | 0.12 | 1.15 (0.82,1.61) | 4.13e-01 | 0.64 (0.53,0.77) | 9.44e-07 |
| rs55786530 | 6 | 32567278 | T | HLA-DRB1 | 0.05 | 0.45 (0.35,0.59) | 9.99e-10 | 0.08 | 1.07 (0.73,1.57) | 7.42e-01 | 0.6 (0.48,0.74) | 9.58e-07 |
| rs17202379 | 6 | 32361449 | A | HCG23 | 0.07 | 0.49 (0.39,0.62) | 2.00e-10 | 0.11 | 1.13 (0.8,1.59) | 4.79e-01 | 0.63 (0.52,0.77) | 9.59e-07 |
| rs9271406 | 6 | 32587588 | G | HLA-DQA1 | 0.35 | 0.62 (0.54,0.72) | 3.38e-11 | 0.41 | 1.13 (0.91,1.41) | 2.64e-01 | 0.74 (0.66,0.84) | 9.78e-07 |
| rs17208741 | 6 | 32361445 | C | HCG23 | 0.07 | 0.49 (0.39,0.62) | 2.08e-10 | 0.11 | 1.13 (0.8,1.59) | 4.79e-01 | 0.63 (0.52,0.77) | 9.85e-07 |
| rs112461335 | 6 | 32548161 | G | HLA-DRB1 | 0.05 | 0.46 (0.35,0.6) | 1.85e-09 | 0.07 | 1.04 (0.69,1.56) | 8.49e-01 | 0.59 (0.47,0.73) | 9.85e-07 |
| rs72850293 | 6 | 32554504 | G | HLA-DRB1 | 0.06 | 0.46 (0.35,0.61) | 5.64e-09 | 0.08 | 0.98 (0.65,1.49) | 9.34e-01 | 0.58 (0.46,0.73) | 9.89e-07 |
| rs28666575 | 6 | 32603944 | T | HLA-DQA1 | 0.05 | 0.46 (0.35,0.6) | 1.44e-09 | 0.09 | 1.05 (0.73,1.5) | 8.00e-01 | 0.61 (0.49,0.76) | 9.91e-07 |
| rs28693734 | 6 | 32603949 | G | HLA-DQA1 | 0.05 | 0.46 (0.35,0.6) | 1.44e-09 | 0.09 | 1.05 (0.73,1.5) | 8.00e-01 | 0.61 (0.49,0.76) | 9.91e-07 |
| rs200431800 | 6 | 32549245 | G | HLA-DRB1 | 0.05 | 0.46 (0.35,0.6) | 1.89e-09 | 0.07 | 1.04 (0.69,1.56) | 8.51e-01 | 0.59 (0.47,0.73) | 9.92e-07 |
| rs72850244 | 6 | 32549638 | A | HLA-DRB1 | 0.05 | 0.46 (0.35,0.6) | 1.87e-09 | 0.07 | 1.04 (0.69,1.56) | 8.47e-01 | 0.59 (0.47,0.73) | 9.99e-07 |
| 6 | 6 | 32548469 | G | HLA-DRB1 | 0.05 | 0.46 (0.35,0.6) | 1.87e-09 | 0.07 | 1.04 (0.69,1.56) | 8.47e-01 | 0.59 (0.47,0.73) | 9.99e-07 |
| rs3830123 | 6 | 32548646 | T | HLA-DRB1 | 0.05 | 0.46 (0.35,0.6) | 1.87e-09 | 0.07 | 1.04 (0.69,1.56) | 8.47e-01 | 0.59 (0.47,0.73) | 9.99e-07 |
| 6 | 6 | 32549443 | A | HLA-DRB1 | 0.05 | 0.46 (0.35,0.6) | 1.87e-09 | 0.07 | 1.04 (0.69,1.56) | 8.47e-01 | 0.59 (0.47,0.73) | 9.99e-07 |
| rs7451330 | 6 | 32295965 | T | C6orf10 | 0.2 | 1.62 (1.34,1.94) | 2.44e-07 | 0.18 | 1.06 (0.8,1.42) | 6.77e-01 | 1.43 (1.23,1.67) | 6.02e-06 |
| rs3117117 | 6 | 32321272 | C | C6orf10 | 0.2 | 1.62 (1.34,1.94) | 2.44e-07 | 0.18 | 1.06 (0.8,1.42) | 6.78e-01 | 1.43 (1.23,1.67) | 6.02e-06 |
| rs9269178 | 6 | 32447598 | A | HLA-DRB9 | 0.41 | 1.49 (1.29,1.72) | 2.87e-08 | 0.38 | 0.98 (0.79,1.23) | 8.69e-01 | 1.32 (1.17,1.49) | 6.02e-06 |
| rs2395180 | 6 | 32407310 | G | HLA-DRA | 0.22 | 0.69 (0.59,0.81) | 2.04e-06 | 0.23 | 0.88 (0.69,1.12) | 3.01e-01 | 0.74 (0.65,0.85) | 6.04e-06 |
| rs2395179 | 6 | 32407302 | G | HLA-DRA | 0.22 | 0.69 (0.59,0.81) | 2.04e-06 | 0.23 | 0.88 (0.69,1.12) | 3.01e-01 | 0.74 (0.65,0.85) | 6.05e-06 |
| rs9269135 | 6 | 32445030 | T | HLA-DRB9 | 0.41 | 1.49 (1.3,1.72) | 2.50e-08 | 0.38 | 0.98 (0.78,1.22) | 8.39e-01 | 1.32 (1.17,1.49) | 6.07e-06 |
| rs70993836 | 6 | 32420434 | C | HLA-DRB9 | 0.08 | 0.53 (0.43,0.66) | 8.21e-09 | 0.11 | 1.09 (0.78,1.51) | 6.24e-01 | 0.66 (0.55,0.8) | 6.07e-06 |
| rs13204032 | 6 | 32420469 | T | HLA-DRB9 | 0.08 | 0.53 (0.43,0.66) | 8.21e-09 | 0.11 | 1.09 (0.78,1.51) | 6.24e-01 | 0.66 (0.55,0.8) | 6.07e-06 |
| rs3129881 | 6 | 32409484 | T | HLA-DRA | 0.22 | 0.7 (0.6,0.81) | 3.76e-06 | 0.23 | 0.86 (0.68,1.09) | 2.23e-01 | 0.74 (0.65,0.84) | 6.08e-06 |
| rs13336147 | 16 | 72985338 | C | ZFHX3 | 0.23 | 0.76 (0.65,0.89) | 5.31e-04 | 0.24 | 0.7 (0.55,0.88) | 3.10e-03 | 0.74 (0.65,0.84) | 6.09e-06 |
| rs3135002 | 6 | 32668439 | A | MTCO3P1 | 0.27 | 1.58 (1.34,1.85) | 2.83e-08 | 0.23 | 0.98 (0.77,1.25) | 8.63e-01 | 1.36 (1.19,1.56) | 6.10e-06 |
| rs3131376 | 6 | 31646683 | A | LY6G5C | 0.19 | 1.51 (1.26,1.83) | 1.20e-05 | 0.18 | 1.27 (0.94,1.71) | 1.12e-01 | 1.44 (1.23,1.69) | 6.10e-06 |
| rs9269176 | 6 | 32447545 | T | HLA-DRB9 | 0.41 | 1.49 (1.29,1.72) | 2.80e-08 | 0.38 | 0.98 (0.78,1.22) | 8.60e-01 | 1.32 (1.17,1.49) | 6.11e-06 |
| rs56210233 | 6 | 32538754 | T | HLA-DRB1 | 0.05 | 0.42 (0.3,0.58) | 2.53e-08 | 0.07 | 1.05 (0.65,1.7) | 8.39e-01 | 0.56 (0.43,0.73) | 6.12e-06 |
| rs55888650 | 6 | 32538756 | G | HLA-DRB1 | 0.05 | 0.42 (0.3,0.58) | 2.53e-08 | 0.07 | 1.05 (0.65,1.7) | 8.39e-01 | 0.56 (0.43,0.73) | 6.12e-06 |
| rs13204265 | 6 | 32420502 | C | HLA-DRB9 | 0.08 | 0.53 (0.42,0.66) | 9.34e-09 | 0.11 | 1.08 (0.78,1.51) | 6.45e-01 | 0.66 (0.55,0.8) | 6.13e-06 |
| rs28895057 | 6 | 32417244 | G | CMAHP | 0.09 | 0.54 (0.43,0.67) | 1.47e-08 | 0.11 | 1.06 (0.76,1.47) | 7.29e-01 | 0.66 (0.55,0.8) | 6.13e-06 |
| rs199504792 | 6 | 32924414 | CT | HLA-DMA | 0.07 | 0.59 (0.46,0.75) | 7.92e-06 | 0.09 | 0.78 (0.55,1.09) | 1.47e-01 | 0.64 (0.53,0.78) | 6.14e-06 |
| rs28895088 | 6 | 32418396 | A | CMAHP | 0.08 | 0.53 (0.43,0.66) | 8.37e-09 | 0.11 | 1.09 (0.78,1.51) | 6.25e-01 | 0.66 (0.55,0.8) | 6.14e-06 |
| rs28895089 | 6 | 32418434 | T | CMAHP | 0.08 | 0.53 (0.43,0.66) | 8.37e-09 | 0.11 | 1.09 (0.78,1.51) | 6.25e-01 | 0.66 (0.55,0.8) | 6.14e-06 |
| rs3115668 | 6 | 31641485 | T | LY6G5B | 0.19 | 1.51 (1.26,1.82) | 1.22e-05 | 0.18 | 1.27 (0.94,1.71) | 1.12e-01 | 1.44 (1.23,1.69) | 6.19e-06 |
| rs9269232 | 6 | 32451760 | T | HLA-DRB9 | 0.31 | 0.74 (0.64,0.85) | 3.16e-05 | 0.34 | 0.81 (0.65,1.01) | 5.68e-02 | 0.76 (0.67,0.86) | 6.20e-06 |
| rs3031757 | 3 | 66309268 | C | SLC25A26 | 0.15 | 1.53 (1.24,1.88) | 7.72e-05 | 0.17 | 1.41 (1.04,1.92) | 2.66e-02 | 1.49 (1.25,1.77) | 6.22e-06 |
| 6 | 6 | 32353620 | T | HCG23 | 0.07 | 0.51 (0.4,0.64) | 2.83e-09 | 0.11 | 1.15 (0.81,1.63) | 4.42e-01 | 0.65 (0.54,0.79) | 6.29e-06 |
| rs78439249 | 5 | 28099919 | A | AC010455.1 | 0.01 | 0.28 (0.14,0.58) | 8.95e-05 | 0.01 | 0.36 (0.15,0.88) | 2.35e-02 | 0.31 (0.18,0.55) | 6.31e-06 |
| rs34990308 | 6 | 32416441 | T | CMAHP | 0.08 | 0.53 (0.43,0.66) | 8.30e-09 | 0.11 | 1.09 (0.78,1.52) | 6.11e-01 | 0.66 (0.55,0.8) | 6.42e-06 |
| rs73926243 | 2 | 46510778 | C | EPAS1 | 0.06 | 1.63 (1.17,2.28) | 3.80e-03 | 0.07 | 3.15 (1.59,6.26) | 1.51e-04 | 1.85 (1.37,2.5) | 6.49e-06 |
| rs112927854 | 6 | 32663259 | T | MTCO3P1 | 0.07 | 0.49 (0.38,0.62) | 2.03e-09 | 0.1 | 1.17 (0.82,1.67) | 3.88e-01 | 0.64 (0.53,0.79) | 6.52e-06 |
| rs35657997 | 6 | 32542236 | G | HLA-DRB1 | 0.06 | 0.49 (0.37,0.64) | 5.95e-08 | 0.07 | 1 (0.64,1.56) | 9.98e-01 | 0.59 (0.47,0.75) | 6.54e-06 |
| rs4868030 | 5 | 1.7E+08 | A | GABRP | 0.17 | 1.49 (1.23,1.79) | 3.32e-05 | 0.17 | 1.33 (0.99,1.8) | 5.74e-02 | 1.44 (1.23,1.69) | 6.56e-06 |
| rs35455030 | 6 | 32522650 | T | HLA-DRB6 | 0.23 | 1.58 (1.32,1.89) | 4.05e-07 | 0.18 | 1.09 (0.79,1.51) | 6.00e-01 | 1.45 (1.24,1.7) | 6.61e-06 |
| rs9273076 | 6 | 32612301 | T | HLA-DQA1 | 0.24 | 1.55 (1.3,1.86) | 1.59e-06 | 0.2 | 1.15 (0.85,1.55) | 3.58e-01 | 1.43 (1.23,1.67) | 6.69e-06 |
| rs146666669 | 3 | 1.43E+08 | T | SLC9A9 | 0.02 | 2.88 (1.53,5.43) | 5.17e-04 | 0.02 | 9.05 (0.72,114.24) | 3.61e-03 | 3.08 (1.67,5.7) | 6.70e-06 |
| rs3134969 | 6 | 32674027 | C | MTCO3P1 | 0.27 | 1.57 (1.34,1.85) | 2.95e-08 | 0.23 | 0.98 (0.76,1.24) | 8.42e-01 | 1.36 (1.19,1.56) | 6.73e-06 |
| rs3134968 | 6 | 32674059 | A | MTCO3P1 | 0.27 | 1.57 (1.34,1.85) | 2.95e-08 | 0.23 | 0.98 (0.76,1.24) | 8.41e-01 | 1.36 (1.19,1.56) | 6.74e-06 |
| rs9269116 | 6 | 32443458 | T | HLA-DRB9 | 0.41 | 1.51 (1.3,1.74) | 1.81e-08 | 0.37 | 0.96 (0.77,1.2) | 7.41e-01 | 1.32 (1.17,1.49) | 6.80e-06 |
| rs28895150 | 6 | 32422077 | C | HLA-DRB9 | 0.08 | 0.53 (0.42,0.66) | 6.77e-09 | 0.11 | 1.1 (0.79,1.54) | 5.59e-01 | 0.66 (0.55,0.8) | 6.83e-06 |
| 6 | 6 | 32422078 | C | HLA-DRB9 | 0.08 | 0.53 (0.42,0.66) | 6.77e-09 | 0.11 | 1.1 (0.79,1.54) | 5.59e-01 | 0.66 (0.55,0.8) | 6.83e-06 |
| 6 | 6 | 32422079 | T | HLA-DRB9 | 0.08 | 0.53 (0.42,0.66) | 6.77e-09 | 0.11 | 1.1 (0.79,1.54) | 5.59e-01 | 0.66 (0.55,0.8) | 6.83e-06 |
| rs61176531 | 6 | 32416058 | C | CMAHP | 0.35 | 0.74 (0.64,0.86) | 8.12e-05 | 0.37 | 0.77 (0.61,0.97) | 2.79e-02 | 0.75 (0.66,0.85) | 6.84e-06 |
| rs28895153 | 6 | 32422086 | T | HLA-DRB9 | 0.08 | 0.53 (0.42,0.66) | 6.80e-09 | 0.11 | 1.1 (0.79,1.54) | 5.60e-01 | 0.66 (0.55,0.8) | 6.85e-06 |
| rs28539606 | 6 | 32626574 | G | CMAHP | 0.08 | 0.52 (0.42,0.65) | 1.90e-09 | 0.13 | 1.16 (0.84,1.6) | 3.69e-01 | 0.67 (0.56,0.81) | 6.85e-06 |
| rs9269503 | 6 | 32543161 | T | HLA-DRB1 | 0.19 | 1.6 (1.31,1.97) | 4.58e-06 | 0.15 | 1.26 (0.87,1.83) | 2.17e-01 | 1.52 (1.27,1.82) | 6.87e-06 |
| rs34491087 | 6 | 32553706 | C | HLA-DRB1 | 0.05 | 0.46 (0.35,0.61) | 9.24e-09 | 0.07 | 1.12 (0.73,1.71) | 6.09e-01 | 0.6 (0.47,0.76) | 6.94e-06 |
| rs9269163 | 6 | 32446833 | A | HLA-DRB9 | 0.41 | 1.5 (1.3,1.73) | 2.03e-08 | 0.37 | 0.97 (0.77,1.21) | 7.57e-01 | 1.32 (1.17,1.49) | 6.95e-06 |
| rs9269165 | 6 | 32446843 | A | HLA-DRB9 | 0.41 | 1.5 (1.3,1.73) | 2.03e-08 | 0.37 | 0.97 (0.77,1.21) | 7.57e-01 | 1.32 (1.17,1.49) | 6.95e-06 |
| rs111461278 | 6 | 32553931 | G | HLA-DRB1 | 0.06 | 0.46 (0.35,0.61) | 6.17e-09 | 0.08 | 1.14 (0.75,1.75) | 5.38e-01 | 0.6 (0.48,0.76) | 6.98e-06 |
| rs10043047 | 5 | 1.7E+08 | G | GABRP | 0.18 | 1.48 (1.23,1.79) | 3.51e-05 | 0.17 | 1.33 (0.99,1.8) | 5.80e-02 | 1.44 (1.23,1.69) | 6.99e-06 |
| rs10039477 | 5 | 1.7E+08 | T | GABRP | 0.18 | 1.48 (1.23,1.79) | 3.53e-05 | 0.17 | 1.33 (0.99,1.8) | 5.80e-02 | 1.44 (1.23,1.69) | 7.02e-06 |
| rs28838605 | 6 | 32510279 | C | RNU1-61P | 0.39 | 0.65 (0.55,0.75) | 1.69e-08 | 0.44 | 1.05 (0.82,1.34) | 7.17e-01 | 0.74 (0.65,0.84) | 7.04e-06 |
| rs9273500 | 6 | 32628342 | T | HLA-DQB1-AS1 | 0.25 | 1.58 (1.33,1.87) | 1.01e-07 | 0.22 | 1.01 (0.78,1.31) | 9.12e-01 | 1.38 (1.2,1.59) | 7.07e-06 |
| rs12191360 | 6 | 32451361 | C | HLA-DRB9 | 0.42 | 1.48 (1.28,1.71) | 4.39e-08 | 0.39 | 0.99 (0.79,1.23) | 9.09e-01 | 1.32 (1.17,1.48) | 7.09e-06 |
| rs9270923 | 6 | 32572405 | C | HLA-DRB1 | 0.22 | 1.5 (1.26,1.79) | 5.06e-06 | 0.21 | 1.19 (0.9,1.57) | 2.10e-01 | 1.4 (1.21,1.63) | 7.11e-06 |
| rs66781242 | 6 | 32507812 | G | HLA-DRB5 | 0.21 | 1.61 (1.35,1.93) | 1.52e-07 | 0.18 | 1.03 (0.77,1.4) | 8.27e-01 | 1.43 (1.23,1.67) | 7.13e-06 |
| rs732292 | 1 | 1.76E+08 | A | RP11-222A5.1 | 0.4 | 1.34 (1.17,1.54) | 2.99e-05 | 0.4 | 1.23 (0.98,1.53) | 6.69e-02 | 1.31 (1.16,1.47) | 7.14e-06 |
| rs743862 | 6 | 32381939 | C | CMAHP | 0.26 | 0.71 (0.62,0.83) | 5.20e-06 | 0.27 | 0.86 (0.69,1.09) | 2.08e-01 | 0.75 (0.67,0.85) | 7.17e-06 |
| rs11377479 | 6 | 32409188 | G | HLA-DRA | 0.22 | 0.7 (0.6,0.81) | 3.76e-06 | 0.23 | 0.87 (0.68,1.1) | 2.48e-01 | 0.74 (0.65,0.85) | 7.17e-06 |
| rs78846716 | 6 | 32553309 | G | HLA-DRB1 | 0.05 | 0.46 (0.35,0.61) | 1.33e-08 | 0.07 | 1.1 (0.72,1.68) | 6.67e-01 | 0.6 (0.47,0.76) | 7.17e-06 |
| rs10404530 | 19 | 52990568 | T | ZNF578 | 0.13 | 1.73 (1.33,2.25) | 3.13e-05 | 0.11 | 1.56 (0.95,2.55) | 6.50e-02 | 1.69 (1.34,2.13) | 7.18e-06 |
| exm-rs9274407 | 6 | 32632832 | T | HLA-DQB1 | 0.22 | 1.56 (1.31,1.86) | 4.35e-07 | 0.19 | 1.07 (0.82,1.4) | 6.09e-01 | 1.4 (1.21,1.62) | 7.21e-06 |
| rs3129874 | 6 | 32407440 | C | HLA-DRA | 0.22 | 0.69 (0.59,0.81) | 2.39e-06 | 0.23 | 0.88 (0.69,1.12) | 3.12e-01 | 0.74 (0.65,0.85) | 7.26e-06 |
| rs3129873 | 6 | 32407433 | C | HLA-DRA | 0.22 | 0.69 (0.59,0.81) | 2.39e-06 | 0.23 | 0.88 (0.69,1.12) | 3.12e-01 | 0.74 (0.65,0.85) | 7.26e-06 |
| rs16870192 | 6 | 32498804 | C | HLA-DRB5 | 0.05 | 0.45 (0.33,0.62) | 1.01e-07 | 0.08 | 0.98 (0.63,1.52) | 9.23e-01 | 0.58 (0.45,0.75) | 7.31e-06 |
| rs55651120 | 6 | 32619835 | T | HLA-DQA1 | 0.08 | 0.52 (0.41,0.67) | 6.76e-08 | 0.15 | 1 (0.72,1.39) | 9.89e-01 | 0.66 (0.54,0.8) | 7.35e-06 |
| rs9271874 | 6 | 32595424 | T | HLA-DQA1 | 0.23 | 1.57 (1.31,1.88) | 6.65e-07 | 0.2 | 1.1 (0.82,1.46) | 5.32e-01 | 1.42 (1.22,1.65) | 7.36e-06 |
| rs67408570 | 6 | 32508874 | C | RNU1-61P | 0.39 | 0.64 (0.54,0.74) | 7.00e-09 | 0.43 | 1.08 (0.84,1.39) | 5.45e-01 | 0.73 (0.64,0.84) | 7.41e-06 |
| rs34691324 | 6 | 32554050 | G | HLA-DRB1 | 0.06 | 0.47 (0.36,0.62) | 1.16e-08 | 0.09 | 1.1 (0.74,1.65) | 6.32e-01 | 0.61 (0.49,0.77) | 7.42e-06 |
| rs113609412 | 6 | 32554047 | A | HLA-DRB1 | 0.06 | 0.47 (0.36,0.62) | 1.16e-08 | 0.09 | 1.1 (0.74,1.65) | 6.32e-01 | 0.61 (0.49,0.77) | 7.42e-06 |
| rs200277885 | 6 | 32359513 | G | HCG23 | 0.08 | 0.52 (0.41,0.65) | 4.63e-09 | 0.11 | 1.13 (0.8,1.59) | 4.77e-01 | 0.66 (0.55,0.8) | 7.44e-06 |
| rs115690055 | 6 | 32359521 | T | HCG23 | 0.08 | 0.52 (0.41,0.65) | 4.63e-09 | 0.11 | 1.13 (0.8,1.59) | 4.77e-01 | 0.66 (0.55,0.8) | 7.44e-06 |
| rs9272777 | 6 | 32610281 | T | HLA-DQA1 | 0.37 | 0.68 (0.59,0.79) | 2.82e-07 | 0.39 | 0.95 (0.75,1.22) | 7.07e-01 | 0.75 (0.66,0.85) | 7.45e-06 |
| rs9270656 | 6 | 32566011 | C | HLA-DRB1 | 0.21 | 1.52 (1.27,1.81) | 2.91e-06 | 0.2 | 1.16 (0.88,1.53) | 2.89e-01 | 1.41 (1.21,1.63) | 7.48e-06 |
| rs201574337 | 6 | 32453496 | C | HLA-DRB9 | 0.36 | 0.62 (0.54,0.72) | 5.43e-11 | 0.42 | 1.23 (0.98,1.54) | 7.82e-02 | 0.76 (0.67,0.85) | 7.48e-06 |
| rs112401921 | 6 | 32487256 | A | HLA-DRB5 | 0.04 | 0.34 (0.22,0.51) | 1.58e-08 | 0.08 | 1.09 (0.71,1.7) | 6.88e-01 | 0.59 (0.44,0.8) | 7.48e-06 |
| rs146394066 | 6 | 32435892 | TA | HLA-DRB9 | 0.06 | 0.47 (0.36,0.61) | 2.52e-09 | 0.08 | 1.19 (0.8,1.76) | 3.88e-01 | 0.62 (0.5,0.77) | 7.49e-06 |
| rs3129306 | 6 | 32953982 | A | CMAHP | 0.07 | 0.68 (0.54,0.87) | 1.83e-03 | 0.07 | 0.53 (0.36,0.76) | 6.79e-04 | 0.63 (0.52,0.77) | 7.52e-06 |
| rs3129876 | 6 | 32408012 | A | HLA-DRA | 0.22 | 0.7 (0.6,0.81) | 2.71e-06 | 0.23 | 0.88 (0.7,1.12) | 3.02e-01 | 0.75 (0.66,0.85) | 7.61e-06 |
| rs13220326 | 6 | 32423602 | A | HLA-DRB9 | 0.08 | 0.54 (0.43,0.67) | 1.44e-08 | 0.11 | 1.08 (0.77,1.5) | 6.62e-01 | 0.66 (0.55,0.8) | 7.67e-06 |
| rs7748270 | 6 | 32448599 | T | HLA-DRB9 | 0.41 | 0.66 (0.57,0.76) | 1.17e-08 | 0.47 | 1.06 (0.84,1.34) | 6.23e-01 | 0.75 (0.67,0.85) | 7.69e-06 |
| rs7791656 | 7 | 47211452 | T | AC004870.3 | 0.32 | 1.41 (1.21,1.65) | 8.06e-06 | 0.3 | 1.18 (0.93,1.5) | 1.71e-01 | 1.34 (1.18,1.53) | 7.70e-06 |
| 6 | 6 | 32594850 | T | HLA-DQA1 | 0.3 | 1.56 (1.32,1.84) | 1.18e-07 | 0.3 | 1.02 (0.79,1.31) | 9.08e-01 | 1.37 (1.19,1.57) | 7.78e-06 |
| rs143440435 | 11 | 32929494 | G | QSER1 | 0.01 | 0.21 (0.08,0.58) | 8.69e-05 | 0.01 | 0.35 (0.13,0.92) | 3.00e-02 | 0.28 (0.14,0.55) | 7.86e-06 |
| rs7776032 | 6 | 32444896 | C | HLA-DRB9 | 0.41 | 1.49 (1.29,1.72) | 4.15e-08 | 0.39 | 0.98 (0.78,1.23) | 8.63e-01 | 1.32 (1.17,1.49) | 7.93e-06 |
| rs7197 | 6 | 32412580 | T | HLA-DRA | 0.24 | 1.52 (1.29,1.8) | 9.91e-07 | 0.21 | 1.1 (0.85,1.42) | 4.78e-01 | 1.38 (1.2,1.59) | 8.02e-06 |
| rs13217272 | 6 | 32423540 | T | HLA-DRB9 | 0.08 | 0.53 (0.43,0.67) | 1.52e-08 | 0.11 | 1.08 (0.77,1.5) | 6.60e-01 | 0.66 (0.55,0.8) | 8.02e-06 |
| rs28895186 | 6 | 32423526 | C | HLA-DRB9 | 0.08 | 0.53 (0.43,0.67) | 1.52e-08 | 0.11 | 1.08 (0.77,1.5) | 6.60e-01 | 0.66 (0.55,0.8) | 8.02e-06 |
| rs9268641 | 6 | 32406887 | T | HLA-DRA | 0.22 | 0.69 (0.59,0.81) | 2.60e-06 | 0.23 | 0.88 (0.7,1.13) | 3.19e-01 | 0.74 (0.65,0.85) | 8.06e-06 |
| rs9272778 | 6 | 32610282 | T | HLA-DQA1 | 0.48 | 1.46 (1.26,1.68) | 1.98e-07 | 0.42 | 1.03 (0.82,1.29) | 8.06e-01 | 1.32 (1.17,1.49) | 8.07e-06 |
| 6 | 6 | 32452602 | T | HLA-DRB9 | 0.34 | 0.63 (0.55,0.73) | 1.28e-10 | 0.39 | 1.2 (0.96,1.5) | 1.10e-01 | 0.76 (0.67,0.86) | 8.12e-06 |
| rs201057798 | 6 | 32359514 | A | HCG23 | 0.07 | 0.52 (0.41,0.65) | 8.01e-09 | 0.11 | 1.11 (0.79,1.57) | 5.42e-01 | 0.66 (0.54,0.8) | 8.17e-06 |
| rs7745797 | 6 | 32550272 | A | HLA-DRB1 | 0.2 | 1.55 (1.28,1.88) | 8.94e-06 | 0.18 | 1.26 (0.9,1.76) | 1.69e-01 | 1.47 (1.24,1.74) | 8.23e-06 |
| rs3117116 | 6 | 32367017 | G | BTNL2 | 0.2 | 1.58 (1.31,1.89) | 7.60e-07 | 0.17 | 1.1 (0.82,1.46) | 5.35e-01 | 1.42 (1.22,1.66) | 8.24e-06 |
| rs140448261 | 2 | 46518723 | C | EPAS1 | 0.06 | 1.65 (1.18,2.31) | 3.03e-03 | 0.07 | 2.82 (1.48,5.34) | 3.33e-04 | 1.85 (1.38,2.49) | 8.28e-06 |
| rs332350 | 3 | 66307348 | A | SLC25A26 | 0.13 | 1.53 (1.24,1.9) | 8.53e-05 | 0.15 | 1.41 (1.02,1.95) | 3.24e-02 | 1.49 (1.25,1.79) | 8.35e-06 |
| rs35092298 | 6 | 32553729 | T | HLA-DRB1 | 0.05 | 0.46 (0.35,0.61) | 1.21e-08 | 0.07 | 1.12 (0.73,1.71) | 6.07e-01 | 0.6 (0.48,0.76) | 8.36e-06 |
| rs28895051 | 6 | 32416909 | G | CMAHP | 0.09 | 0.53 (0.43,0.67) | 9.10e-09 | 0.12 | 1.1 (0.79,1.54) | 5.56e-01 | 0.67 (0.56,0.8) | 8.40e-06 |
| rs28895192 | 6 | 32424943 | G | HLA-DRB9 | 0.08 | 0.53 (0.42,0.66) | 8.89e-09 | 0.11 | 1.11 (0.79,1.55) | 5.50e-01 | 0.66 (0.55,0.8) | 8.47e-06 |
| rs13220311 | 6 | 32423563 | A | HLA-DRB9 | 0.08 | 0.54 (0.43,0.67) | 1.68e-08 | 0.11 | 1.08 (0.77,1.5) | 6.62e-01 | 0.67 (0.55,0.8) | 8.51e-06 |
| rs28895145 | 6 | 32421861 | C | HLA-DRB9 | 0.09 | 0.53 (0.43,0.67) | 8.91e-09 | 0.12 | 1.11 (0.79,1.54) | 5.49e-01 | 0.67 (0.56,0.8) | 8.51e-06 |
| rs9269126 | 6 | 32444915 | C | HLA-DRB9 | 0.41 | 1.49 (1.29,1.72) | 4.53e-08 | 0.39 | 0.98 (0.78,1.23) | 8.59e-01 | 1.32 (1.17,1.49) | 8.54e-06 |
| rs61735550 | 16 | 72992221 | A | ZFHX3 | 0.23 | 0.75 (0.64,0.88) | 3.09e-04 | 0.24 | 0.72 (0.57,0.92) | 8.99e-03 | 0.74 (0.65,0.85) | 8.55e-06 |
| rs185608970 | 10 | 97292877 | T | SORBS1 | 0.01 | 12.28 (0.85,176.93) | 2.64e-04 | 0.02 | 5.89 (0.98,35.5) | 1.08e-02 | 7.41 (1.67,32.86) | 8.62e-06 |
| rs71691419 | 6 | 1.59E+08 | T | CACYBPP3 | 0.04 | 0.63 (0.47,0.86) | 3.32e-03 | 0.05 | 0.45 (0.3,0.7) | 3.01e-04 | 0.57 (0.44,0.73) | 8.64e-06 |
| rs17202717 | 6 | 32397863 | T | HLA-DRA | 0.06 | 0.46 (0.36,0.6) | 8.77e-10 | 0.09 | 1.26 (0.85,1.84) | 2.40e-01 | 0.63 (0.51,0.78) | 8.65e-06 |
| rs17496307 | 6 | 32401036 | T | HLA-DRA | 0.06 | 0.46 (0.36,0.6) | 8.77e-10 | 0.09 | 1.26 (0.85,1.84) | 2.40e-01 | 0.63 (0.51,0.78) | 8.65e-06 |
| rs28895179 | 6 | 32423294 | G | HLA-DRB9 | 0.08 | 0.54 (0.43,0.67) | 1.46e-08 | 0.11 | 1.08 (0.78,1.51) | 6.32e-01 | 0.67 (0.55,0.8) | 8.65e-06 |
| rs28895180 | 6 | 32423298 | T | HLA-DRB9 | 0.08 | 0.54 (0.43,0.67) | 1.46e-08 | 0.11 | 1.08 (0.78,1.51) | 6.32e-01 | 0.67 (0.55,0.8) | 8.65e-06 |
| rs3130068 | 6 | 31590354 | G | PRRC2A | 0.18 | 1.52 (1.25,1.83) | 1.71e-05 | 0.18 | 1.27 (0.94,1.72) | 1.16e-01 | 1.44 (1.23,1.7) | 8.66e-06 |
| rs6124391 | 20 | 40530954 | C | RP5-1121H13.4 | 0.15 | 1.46 (1.19,1.79) | 2.23e-04 | 0.13 | 1.55 (1.09,2.22) | 1.32e-02 | 1.48 (1.24,1.77) | 8.71e-06 |
| rs73926244 | 2 | 46510792 | G | EPAS1 | 0.06 | 1.66 (1.18,2.32) | 3.01e-03 | 0.06 | 3.11 (1.51,6.4) | 3.64e-04 | 1.86 (1.37,2.52) | 8.75e-06 |
| rs9271147 | 6 | 32577385 | T | HLA-DQA1 | 0.21 | 1.52 (1.27,1.81) | 3.37e-06 | 0.19 | 1.16 (0.88,1.53) | 2.97e-01 | 1.41 (1.21,1.63) | 8.78e-06 |
| rs10527169 | 6 | 31588984 | CGTG | PRRC2A | 0.22 | 1.44 (1.21,1.71) | 4.43e-05 | 0.22 | 1.3 (0.99,1.71) | 5.92e-02 | 1.4 (1.21,1.62) | 8.81e-06 |
| 4 | 4 | 1.69E+08 | <CN0> | PHBP14 | 0.01 | 0.24 (0.09,0.64) | 3.86e-04 | 0.01 | 0.21 (0.06,0.72) | 7.20e-03 | 0.23 (0.11,0.49) | 8.82e-06 |
| rs906734 | 12 | 1.28E+08 | G | RP11-955H22.1 | 0.18 | 0.76 (0.64,0.91) | 2.16e-03 | 0.17 | 0.62 (0.47,0.81) | 6.50e-04 | 0.72 (0.62,0.83) | 8.83e-06 |
| rs9268632 | 6 | 32406412 | G | HLA-DRA | 0.22 | 0.7 (0.6,0.81) | 2.95e-06 | 0.23 | 0.88 (0.7,1.12) | 3.18e-01 | 0.75 (0.65,0.85) | 8.86e-06 |
| rs28895193 | 6 | 32424954 | A | HLA-DRB9 | 0.08 | 0.53 (0.42,0.66) | 8.98e-09 | 0.11 | 1.11 (0.79,1.55) | 5.39e-01 | 0.66 (0.55,0.8) | 8.89e-06 |
| rs13203280 | 6 | 32449961 | T | HLA-DRB9 | 0.05 | 0.45 (0.34,0.59) | 2.29e-09 | 0.08 | 1.21 (0.81,1.81) | 3.40e-01 | 0.62 (0.49,0.77) | 8.91e-06 |
| 6 | 6 | 32444933 | T | HLA-DRB9 | 0.41 | 1.49 (1.29,1.71) | 4.83e-08 | 0.39 | 0.98 (0.78,1.23) | 8.56e-01 | 1.32 (1.17,1.49) | 9.01e-06 |
| rs3135343 | 6 | 32396313 | T | HLA-DRA | 0.22 | 0.7 (0.6,0.81) | 3.35e-06 | 0.23 | 0.88 (0.7,1.12) | 3.02e-01 | 0.75 (0.66,0.85) | 9.02e-06 |
| rs3129845 | 6 | 32396277 | G | HLA-DRA | 0.22 | 0.7 (0.6,0.81) | 3.35e-06 | 0.23 | 0.88 (0.7,1.12) | 3.02e-01 | 0.75 (0.66,0.85) | 9.03e-06 |
| rs13215138 | 6 | 32418923 | G | CMAHP | 0.08 | 0.54 (0.43,0.67) | 1.46e-08 | 0.11 | 1.09 (0.78,1.52) | 6.19e-01 | 0.67 (0.55,0.8) | 9.07e-06 |
| rs71536136 | 6 | 32418954 | TA | CMAHP | 0.08 | 0.54 (0.43,0.67) | 1.46e-08 | 0.11 | 1.09 (0.78,1.52) | 6.19e-01 | 0.67 (0.55,0.8) | 9.07e-06 |
| exm-rs3129900 | 6 | 32305979 | G | C6orf10 | 0.23 | 1.55 (1.3,1.84) | 5.34e-07 | 0.2 | 1.07 (0.81,1.4) | 6.31e-01 | 1.39 (1.2,1.61) | 9.08e-06 |
| rs732291 | 1 | 1.76E+08 | T | RP11-222A5.1 | 0.4 | 1.32 (1.15,1.52) | 7.65e-05 | 0.41 | 1.26 (1.01,1.58) | 3.87e-02 | 1.31 (1.16,1.47) | 9.10e-06 |
| rs13199346 | 6 | 32418986 | T | CMAHP | 0.08 | 0.54 (0.43,0.67) | 1.46e-08 | 0.11 | 1.09 (0.78,1.52) | 6.17e-01 | 0.67 (0.55,0.8) | 9.14e-06 |
| rs13199014 | 6 | 32419012 | C | CMAHP | 0.08 | 0.54 (0.43,0.67) | 1.46e-08 | 0.11 | 1.09 (0.78,1.52) | 6.17e-01 | 0.67 (0.55,0.8) | 9.15e-06 |
| rs13198982 | 6 | 32418893 | T | CMAHP | 0.08 | 0.54 (0.43,0.67) | 1.48e-08 | 0.11 | 1.09 (0.78,1.52) | 6.19e-01 | 0.67 (0.55,0.8) | 9.16e-06 |
| rs28895096 | 6 | 32418859 | C | CMAHP | 0.08 | 0.54 (0.43,0.67) | 1.49e-08 | 0.11 | 1.09 (0.78,1.52) | 6.19e-01 | 0.67 (0.55,0.8) | 9.19e-06 |
| rs28895097 | 6 | 32418869 | T | CMAHP | 0.08 | 0.54 (0.43,0.67) | 1.49e-08 | 0.11 | 1.09 (0.78,1.52) | 6.19e-01 | 0.67 (0.55,0.8) | 9.19e-06 |
| rs13215109 | 6 | 32418880 | C | CMAHP | 0.08 | 0.54 (0.43,0.67) | 1.49e-08 | 0.11 | 1.09 (0.78,1.52) | 6.19e-01 | 0.67 (0.55,0.8) | 9.19e-06 |
| rs71534537 | 6 | 32511486 | A | RNU1-61P | 0.28 | 0.61 (0.51,0.72) | 4.14e-09 | 0.34 | 1.12 (0.86,1.45) | 4.11e-01 | 0.73 (0.63,0.84) | 9.24e-06 |
| rs111371469 | 6 | 32511491 | G | RNU1-61P | 0.28 | 0.61 (0.51,0.72) | 4.14e-09 | 0.34 | 1.12 (0.86,1.45) | 4.11e-01 | 0.73 (0.63,0.84) | 9.24e-06 |
| rs1007636 | 6 | 32904041 | C | HLA-DMB | 0.11 | 0.74 (0.6,0.91) | 4.67e-03 | 0.09 | 0.51 (0.36,0.73) | 1.76e-04 | 0.67 (0.56,0.8) | 9.24e-06 |
| rs17208671 | 6 | 32360849 | T | HCG23 | 0.13 | 0.57 (0.47,0.68) | 3.80e-10 | 0.17 | 1.23 (0.92,1.65) | 1.62e-01 | 0.7 (0.6,0.82) | 9.26e-06 |
| rs4476870 | 6 | 32613417 | G | HLA-DQA1 | 0.06 | 0.47 (0.36,0.61) | 6.95e-09 | 0.1 | 1.14 (0.79,1.63) | 4.87e-01 | 0.64 (0.52,0.79) | 9.28e-06 |
| rs9268149 | 6 | 32263099 | T | C6orf10 | 0.23 | 1.52 (1.29,1.81) | 9.54e-07 | 0.21 | 1.09 (0.83,1.43) | 5.22e-01 | 1.39 (1.2,1.6) | 9.31e-06 |
| rs13205804 | 6 | 32663397 | G | MTCO3P1 | 0.06 | 0.48 (0.38,0.62) | 5.86e-09 | 0.09 | 1.15 (0.79,1.66) | 4.59e-01 | 0.64 (0.52,0.79) | 9.34e-06 |
| rs76054429 | 19 | 12489670 | C | CTD-3105H18.14 | 0.01 | 0.2 (0.07,0.54) | 9.83e-05 | 0.01 | 0.31 (0.1,0.96) | 3.18e-02 | 0.24 (0.11,0.51) | 9.35e-06 |
| rs72847642 | 6 | 32542071 | T | HLA-DRB1 | 0.06 | 0.5 (0.39,0.65) | 5.46e-08 | 0.08 | 1.04 (0.67,1.61) | 8.70e-01 | 0.61 (0.48,0.76) | 9.37e-06 |
| rs112938446 | 6 | 32536893 | C | CMAHP | 0.05 | 0.41 (0.3,0.55) | 4.65e-10 | 0.11 | 1.31 (0.88,1.94) | 1.74e-01 | 0.63 (0.49,0.8) | 9.42e-06 |
| rs9271323 | 6 | 32582454 | G | HLA-DQA1 | 0.1 | 0.64 (0.52,0.78) | 1.07e-05 | 0.12 | 0.81 (0.6,1.09) | 1.67e-01 | 0.69 (0.58,0.81) | 9.46e-06 |
| rs28507026 | 6 | 32582485 | G | HLA-DQA1 | 0.1 | 0.64 (0.52,0.78) | 1.07e-05 | 0.12 | 0.81 (0.6,1.09) | 1.67e-01 | 0.69 (0.58,0.81) | 9.46e-06 |
| rs10773380 | 12 | 1.28E+08 | T | RP11-955H22.1 | 0.19 | 0.75 (0.64,0.89) | 8.20e-04 | 0.18 | 0.67 (0.52,0.87) | 3.05e-03 | 0.73 (0.63,0.84) | 9.54e-06 |
| rs35703034 | 6 | 32555124 | C | HLA-DRB1 | 0.06 | 0.46 (0.35,0.6) | 3.80e-09 | 0.1 | 1.18 (0.81,1.73) | 3.90e-01 | 0.63 (0.51,0.79) | 9.60e-06 |
| rs3129870 | 6 | 32406100 | T | HLA-DRA | 0.22 | 0.7 (0.6,0.81) | 3.29e-06 | 0.23 | 0.88 (0.7,1.12) | 3.17e-01 | 0.75 (0.66,0.85) | 9.65e-06 |
| rs35817460 | 6 | 32394122 | T | HLA-DRA | 0.06 | 0.46 (0.36,0.6) | 1.07e-09 | 0.09 | 1.25 (0.85,1.84) | 2.41e-01 | 0.63 (0.51,0.78) | 9.69e-06 |
| rs9469112 | 6 | 32415153 | T | CMAHP | 0.1 | 0.57 (0.47,0.7) | 6.15e-08 | 0.13 | 1.02 (0.75,1.4) | 8.84e-01 | 0.68 (0.58,0.81) | 9.72e-06 |
| rs146605120 | 21 | 18307451 | C | AF212831.2 | 0.01 | 0.32 (0.16,0.64) | 3.31e-04 | 0.01 | 0.32 (0.13,0.78) | 9.60e-03 | 0.32 (0.19,0.56) | 9.72e-06 |
| rs9272364 | 6 | 32604599 | A | HLA-DQA1 | 0.21 | 0.61 (0.52,0.71) | 7.04e-10 | 0.25 | 1.18 (0.91,1.52) | 2.02e-01 | 0.74 (0.64,0.84) | 9.82e-06 |
| rs9281806 | 6 | 32403240 | GT | HLA-DRA | 0.22 | 0.7 (0.6,0.81) | 3.39e-06 | 0.23 | 0.88 (0.7,1.12) | 3.17e-01 | 0.75 (0.66,0.85) | 9.87e-06 |
| rs73397431 | 6 | 32523385 | A | HLA-DRB6 | 0.24 | 1.53 (1.29,1.82) | 1.24e-06 | 0.19 | 1.12 (0.81,1.54) | 4.87e-01 | 1.42 (1.22,1.66) | 9.88e-06 |
| rs200933141 | 16 | 88115009 | TAAAAAAAAA | RP11-863P13.5 | 0.16 | 1.58 (1.26,1.97) | 4.84e-05 | 0.15 | 1.39 (0.98,1.97) | 6.11e-02 | 1.52 (1.26,1.84) | 9.89e-06 |
| rs146329501 | 5 | 1.7E+08 | T | GABRP | 0.17 | 1.49 (1.23,1.8) | 3.23e-05 | 0.17 | 1.3 (0.96,1.76) | 8.34e-02 | 1.43 (1.22,1.68) | 9.94e-06 |
| rs28895094 | 6 | 32418843 | T | CMAHP | 0.08 | 0.54 (0.43,0.67) | 1.72e-08 | 0.11 | 1.09 (0.78,1.51) | 6.23e-01 | 0.67 (0.56,0.8) | 9.95e-06 |
| rs3135398 | 6 | 32403934 | T | HLA-DRA | 0.22 | 0.7 (0.6,0.81) | 3.42e-06 | 0.23 | 0.88 (0.7,1.12) | 3.18e-01 | 0.75 (0.66,0.85) | 9.96e-06 |
| rs3129864 | 6 | 32404043 | G | HLA-DRA | 0.22 | 0.7 (0.6,0.81) | 3.43e-06 | 0.23 | 0.88 (0.7,1.12) | 3.18e-01 | 0.75 (0.66,0.85) | 9.98e-06 |
| rs28895123 | 6 | 32420828 | C | HLA-DRB9 | 0.08 | 0.52 (0.42,0.66) | 7.52e-09 | 0.11 | 1.13 (0.8,1.59) | 4.82e-01 | 0.66 (0.55,0.8) | 9.98e-06 |
| SNP: single nucleotide polymorphism; Chr: chromosome; MAF: minor allele frequency; OR: odds ratio; 95% CI: 95% confidence interval | | | | | | | | | | | | |

| **Table S4**. Odds ratios and p-values for candidate genes | | | | | | | | | | | | |
| --- | --- | --- | --- | --- | --- | --- | --- | --- | --- | --- | --- | --- |
|  | | | | | Phase1 | | | Phase 2 | | | Meta-analysis | |
| SNP | Chr | position | Minor allele | Gene | MAF case | OR (95% CI) | p-value | MAF case | OR (95% CI) | p-value | OR (95% CI) | p-value |
| rs715299 | 6 | 32189841 | G | NOTCH4 | 0.31 | 0.96 (0.83,1.11) | 0.56 | 0.29 | 0.90 (0.72,1.14) | 0.39 | 0.94 (0.83,1.07) | 0.34 |
| rs1040461 | 6 | 57055354 | T | RAB23 | 0.08 | 1.13 (0.88,1.44) | 0.35 | 0.09 | 1.16 (0.78,1.72) | 0.46 | 1.14 (0.92,1.40) | 0.23 |
| rs1398024 | 10 | 23665438 | T | C10ORF67 | 0.23 | 1.20 (1.02,1.41) | 0.02 | 0.24 | 0.97 (0.76,1.25) | 0.83 | 1.13 (0.99,1.29) | 0.08 |
| rs1953600 | 10 | 81911725 | T | ANXA11 | 0.38 | 0.80 (0.70,0.92) | 1.17E-03 | 0.4 | 0.91 (0.73,1.12) | 0.37 | 0.83 (0.74,0.93) | 1.37E-03 |
| rs2573346 | 10 | 81918041 | A | ANXA11 | 0.38 | 0.80 (0.70,0.92) | 1.22E-03 | 0.4 | 0.90 (0.73,1.12) | 0.36 | 0.83 (0.74,0.93) | 1.40E-03 |
| rs2784773 | 10 | 81919800 | T | ANXA11 | 0.41 | 1.20 (1.05,1.37) | 8.49E-03 | 0.38 | 0.96 (0.77,1.19) | 0.70 | 1.13 (1.00,1.26) | 0.05 |
| rs1049550 | 10 | 81926702 | A | ANXA11 | 0.35 | 0.78 (0.68,0.89) | 2.58E-04 | 0.39 | 0.91 (0.73,1.12) | 0.37 | 0.81 (0.73,0.91) | 3.40E-04 |
| rs479777 | 11 | 64107477 | C | CCDC88B | 0.29 | 0.83 (0.72,0.96) | 0.01 | 0.31 | 1.04 (0.82,1.32) | 0.73 | 0.88 (0.78,1.00) | 0.05 |
| rs1050045 | 12 | 58115271 | C | OS9 | 0.46 | 1.18 (1.03,1.35) | 0.02 | 0.47 | 1.13 (0.92,1.39) | 0.25 | 1.16 (1.04,1.30) | 8.63E-03 |
| SNP: single nucleotide polymorphism; Chr: chromosome; MAF: minor allele frequency; OR: odds ratio; 95% CI: 95% confidence interval | | | | | | | | | | | | |

| **Table S5**. Other significant HLA alleles associated with sarcoidosis (p<0.00011 but >5 x 10^-8^) | | | | | | | |
| --- | --- | --- | --- | --- | --- | --- | --- |
|  | | Phase 1 | | Phase 2 | | Meta-analysis | |
| HLA allele | HLA | OR (95% CI) | p-value | OR (95% CI) | p-value | OR (95% CI) | p-value |
| B.35.01 | B | 0.46 (0.32, 0.67) | 3.76E-05 | 0.49 (0.29, 0.83) | 8.50E-03 | 0.47 (0.35, 0.64) | 1.01E-06 |
| DQB1.06.02 | DQB1 | 1.57 (1.31, 1.88) | 1.11E-06 | 1.23 (0.91, 1.66) | 1.86E-01 | 1.47 (1.26, 1.72) | 1.53E-06 |
| DRB1.15.01 | DRB1 | 1.56 (1.30, 1.86) | 1.03E-06 | 1.20 (0.90, 1.61) | 2.12E-01 | 1.45 (1.25, 1.69) | 1.75E-06 |
| DQA1.01.02 | DQA1 | 1.43 (1.23, 1.66) | 4.42E-06 | 1.19 (0.93, 1.54) | 1.66E-01 | 1.36 (1.20, 1.55) | 4.08E-06 |
| DPB1.04.01 | DPB1 | 0.75 (0.66, 0.86) | 4.95E-05 | 0.84 (0.67, 1.06) | 1.36E-01 | 0.78 (0.70, 0.87) | 2.47E-05 |
| DPB1.01.01 | DPB1 | 1.65 (1.29, 2.10) | 5.66E-05 | 1.22 (0.82, 1.83) | 3.24E-01 | 1.52 (1.24, 1.87) | 8.99E-05 |
| SNP: single nucleotide polymorphism; Chr: chromosome; MAF: minor allele frequency; OR: odds ratio; 95% CI: 95% confidence interval | | | | | | | |

| **Table S6.** Summary of top 7 significant SNPs of the genome-wide association study of sarcoidosis adjusted for DRB1*0101, DQA1*0101, and DQB1*0501 (all together) | | | | | | | | | | | | |
| --- | --- | --- | --- | --- | --- | --- | --- | --- | --- | --- | --- | --- |
|  | | | | | Phase1 | | | Phase 2 | | | Meta-analysis | |
| SNP | Chr | position | Minor allele | Nearest gene (+/-25K) | MAF case | OR (95% CI) | p-value | MAF case | OR (95% CI) | p-value | OR (95% CI) | p-value |
| rs9269233 | 6 | 32451762 | A | HLA-DRB9 | 0.38 | 1.60 (1.37, 1.87) | 3.37E-09 | 0.34 | 1.06 (0.83, 1.35) | 0.63 | 1.42 (1.24, 1.61) | 2.15E-07 |
| rs9271346 | 6 | 32583468 | C | HLA-DQA1 | 0.27 | 1.61 (1.36, 1.91) | 3.78E-08 | 0.24 | 1.19 (0.91, 1.54) | 0.20 | 1.47 (1.27, 1.70) | 1.23E-07 |
| rs35656642 | 6 | 32583610 | A | HLA-DQA1 | 0.32 | 0.74 (0.64, 0.87) | 1.24E-04 | 0.34 | 0.84 (0.66, 1.05) | 0.13 | 0.77 (0.68, 0.87) | 5.35E-05 |
| rs28589559 | 6 | 32587716 | T | HLA-DQA1 | 0.08 | 0.63 (0.44, 0.90) | 0.01 | 0.13 | 0.92 (0.68, 1.25) | 0.60 | 0.78 (0.62, 0.99) | 0.02 |
| rs9276935 | 6 | 32936441 | C | BRD2 | 0.05 | 0.50 (0.38, 0.67) | 2.15E-06 | 0.05 | 0.61 (0.40, 0.94) | 0.03 | 0.54 (0.42, 0.68) | 2.21E-07 |
| rs3129888 | 6 | 32411726 | G | HLA-DRA | 0.27 | 1.52 (1.29, 1.79) | 8.46E-07 | 0.25 | 1.18 (0.92, 1.52) | 0.20 | 1.41 (1.22, 1.62) | 1.51E-06 |
| rs71549283 | 6 | 32505038 | A | HLA-DRB5 | 0.33 | 0.69 (0.57, 0.82) | 5.16E-05 | 0.34 | 0.84 (0.63, 1.11) | 0.22 | 0.73 (0.62, 0.85) | 5.00E-05 |
| SNP: single nucleotide polymorphism; Chr: chromosome; MAF: minor allele frequency; OR: odds ratio; 95% CI: 95% confidence interval | | | | | | | | | | | | |

| **Table S7**: Interaction between top 3 SNPs and DRB1*0101* | | | | | | | |
| --- | --- | --- | --- | --- | --- | --- | --- |
| Model 1: sarcoidosis (yes/no) = rs9269233 + DRB1*0101 + DRB1*0101 X rs9269233 + covariates | | | | | | | |
|  | rs9269233 |  |  | DRB1*0101 | DRB1*0101 X rs9269233 |  |  |
| Phase 1 | 2.46E-08 |  |  | 0.39 | 0.57 |  |  |
| Phase 2 | 0.67 |  |  | 0.05 | 0.90 |  |  |
| Meta-analysis | 1.09E-06 |  |  | 0.07 | 0.59 |  |  |
| Model 2: sarcoidosis (yes/no) = rs9271346 + DRB1*0101 + DRB1*0101 X rs9271346 + covariates | | | | | | | |
|  |  | rs9271346 |  | DRB1*0101 |  | DRB1*0101 X rs9271346 |  |
| Phase 1 |  | 1.3E-06 |  | 0.87 |  | 0.11 |  |
| Phase 2 |  | 0.71 |  | 0.79 |  | 0.06 |  |
| Meta-analysis |  | 2.31E-05 |  | 0.99 |  | **0.02** |  |
| Model 3: sarcoidosis (yes/no) = rs35656642 + DRB1*0101 + DRB1*0101 X rs35656642 + covariates | | | | | | | |
|  |  |  | rs35656642 | DRB1*0101 |  |  | DRB1*0101 X rs35656642 |
| Phase 1 |  |  | 6.23E-04 | 0.47 |  |  | 0.30 |
| Phase 2 |  |  | 0.23 | 1.49E-03 |  |  | 0.58 |
| Meta-analysis |  |  | 4.35E-04 | 0.02 |  |  | 0.24 |
| SNP: single nucleotide polymorphism  *presented as p-values; significant p-value<0.05 are bolded | | | | | | | |

| **Table S8**: HLA alleles and their association with sarcoidosis and rheumatoid arthritis in phenome-wide association study | | | | | | |
| --- | --- | --- | --- | --- | --- | --- |
| HLA alleles | MAF | Diagnosis | Cases number | Controls number | OR (95% CI) | p-value |
| DQA1*0101 | 0.14 | Sarcoidosis | 256 | 24376 | 0.74 (0.55,0.99) | 0.05 |
| DQA1*0101 | 0.14 | Rheumatoid arthritis | 1072 | 23403 | 1.36 (1.19,1.55) | 5.92E-06 |
| DQB1*0501 | 0.12 | Sarcoidosis | 256 | 24376 | 0.59 (0.42,0.84) | 3.16E-03 |
| DQB1*0501 | 0.12 | Rheumatoid arthritis | 1072 | 23403 | 1.52 (1.33,1.74) | 1.64E-09 |
| DRB1*0101 | 0.1 | Sarcoidosis | 256 | 24376 | 0.64 (0.44,0.92) | 0.02 |
| DRB1*0101 | 0.1 | Rheumatoid arthritis | 1072 | 23403 | 1.59 (1.38,1.83) | 1.37E-10 |
| MAF: minor allele frequency; OR: odds ratio; 95% CI: 95% confidence interval | | | | | | |

**Figure S1.** Linkage disequilibrium plot of the significant SNPs. The number in the matrix represents the ^r2^ based on CEU population calculated from LD Link (<https://ldlink.nci.nih.gov/>). Top 7 SNPs represented all SNPs (r^2^ >0.70) were circled in red.

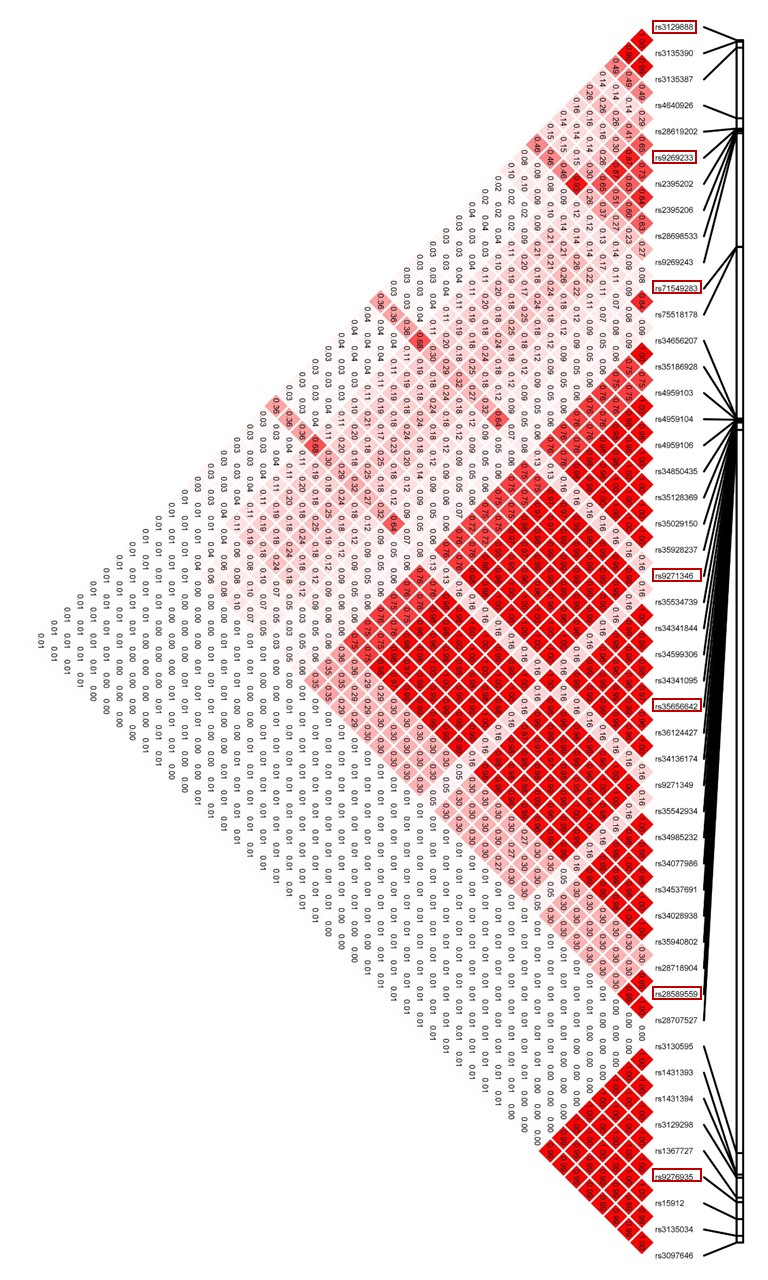

**Figure S2**. Colocolization plot of *HLA-DRA*. Z scores are presented on the Y-axis, with the genomic position on the X-axis. Pairwise linkage disequilibrium (LD) values are shown in the bottom half, and the darker the dot, the higher the LD between the SNPs. rs3135387, rs3129888, and rs3135390 have high GWAS, PBMC, and BAL eQTL Z-scores but have strong LD with each other.

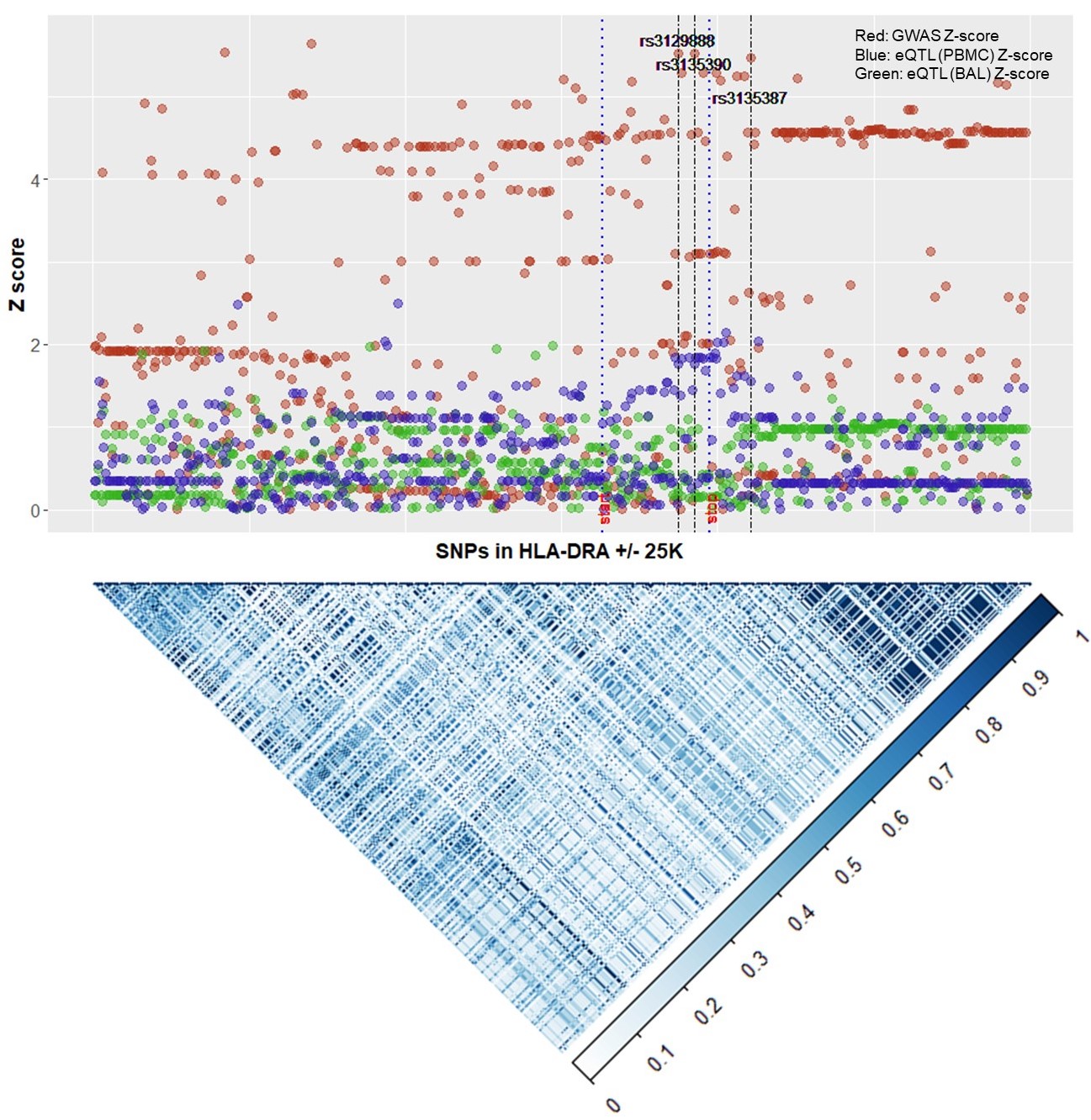
