## Supplemental Method for "Genome-wide association study identifies multiple HLA loci for sarcoidosis susceptibility"

**Supplemental Methods**

**Methods**

Study design and population:

This GWAS used a two-phase approach to identify genome-wide significant SNPs associated with sarcoidosis susceptibility; Phase 2 DNA samples became available after the Phase 1 group had been genotyped. DNA samples from sarcoidosis cases and healthy controls were obtained from established National Jewish Health (NJH) biorepositories as well as enrollment of additional cases at NJH, University of California, San Fransisco (UCSF), Genomic Research in Alpha-1 Antitrypsin Deficiency and Sarcoidosis (GRADS) consortium and Cleveland Clinic Foundation. Additional healthy controls were included from other studies at NJH (Chronic Beryllium Disease Biorepository, Beryllium Biobank, Study of Allergen, Genetics, and Endotoxin), Cleveland Clinic, and UCSF. This study was approved by the Institutional Review Board at NJH (HS2765). Written informed consent was obtained for all participants and all biorepositories. The sarcoidosis case definition was based on the ATS/ERS/WASOG statement in both phases[1]. After quality control (see below), these 818 cases and 981 controls (Phase 1) and 517 cases and 283 controls (Phase 2), all of self-reported European ancestry (**Table 1** and **Figure 1**), reflected the large majority of the race/ethnicities seen in our clinics. We completed duplicate genotyping on approximately 10% of our cohort (n=328) to ensure the reproducibility of genotyping results. Duplicated samples were removed from the final analysis. A subset of study participants in our GWAS had peripheral blood mononuclear cell (PBMC) and/or bronchoalveolar lavage (BAL) cell RNA sequencing data available through the GRADS study[2, 3]. All recruitment and consenting procedures for this population were approved by the institutional review board through GRADS (HS2780). We also tested the significant SNPs identified in our EA two-phase approach in an AA study population. The AA GWAS summary statistics were obtained from a published study[4] with updated imputation since the publication of those data; new imputation of genotypes was completed using the TOPMed imputation panel[5] in order to improve breadth of variant coverage and accuracy compared to the IMPUTE2 default panel (Human Genome).

Genome-wide genotyping:

Subjects in both phases underwent the same protocol for DNA extraction and genotyping. DNA was extracted from whole blood using the PAXgene Blood DNA kit. Genotyping was performed using the Illumina HumanOmini 2.5 BeadChip to interrogate ~ 2.4 million markers. The markers were derived from the 1000 Genomes Project[6], including all three HapMap phases, 19K SNPs across the MHC, and over 41K non-synonymous SNPs. Genotyping of both phases was conducted at Hudson Alpha Biotechnology Institute, a non-profit genomics technologies institution in Huntsville, AL (https://hudsonalpha.org/).

Genotype quality control:

We included only one individual among first-degree relatives based on an estimated kinship coefficient ≥ 0.177 to ensure all subjects were independent of each other. We used the 1000 genomes data[6] on 2,504 individuals (EUR, EAS, AMR, SAS, AFR) as representatives of European, West African, and East Asian populations to infer ancestry-informative principal components (PCs), which were projected onto the case and control samples. We used the first 20 PCs as input and used a support vector machine model (R package "kernlab" with ksvm function )[7] to identify putative non-European samples and removed those samples from all subsequent GWAS analyses (n=311); The EA individuals who also had gene expression data available were included in the expression quantitative trait analyses. In addition to the exclusion of putative non-European samples, we excluded cases and controls with 1) unresolved sex mix-match between clinical and genomic data, 2) genotype calls at less than 98% of SNPs that pass laboratory quality control, and 3) subjects with a diagnosis other than "sarcoidosis" or "control." In addition, we prioritized SNPs for follow-up based on other criteria. We excluded SNPs with differential missingness between cases and controls based on a chi-squared test of proportions of missingness between cases and controls (p-value <0.05). We also tested for departures from Hardy–Weinberg Equilibrium (HWE) via the 1-degree of freedom goodness of fit test. We prioritized SNPs with minor allele frequency (MAF) > 0.03 and HWE p-value > 0.001 in cases and controls evaluated separately, and with < 10% missing data.

Imputation of additional genotypes and HLA variants for the EA population:

We imputed genotypes using the combined case and control discovery samples for all 1000 genomes SNPs. We used the multi-population reference panel data from 1000 genomes for pre-phasing using Shapeit with the default parameters[8, 9]. The imputation was performed using the Impute2 program[10, 11]. To further investigate the HLA region, we imputed classical HLA alleles using the R package HLA Genotype Imputation with Attribute Bagging (HIBAG)[12]. The average posterior probability was derived from HIBAG using the ensemble classifier and bagging techniques. We used the EA reference panel for the imputation.

RNA sequencing and quality control:

Total RNA was extracted, and RNA-sequencing was conducted as outlined in the recent GRADS publication[2]. The alignment was based onGRCh37. We followed a similar quality control procedure for both PBMC and BAL samples and removed RNA samples with an unmapped read rate > 20% and mitochondrial read rate >10%. We also removed outlier samples through PC analysis.

Statistical analysis:

*Single-SNP association test and meta-analysis*

We tested for association between each SNP and sarcoidosis using Snptest (v2)[13] as described previously[14]. To obtain an overall measure of association with sarcoidosis, we performed a meta-analysis of Phase 1 and Phase 2 using summary statistic data and the weighted inverse normal method[15] as implemented in the software METAL[16]. Genome-wide significance was defined as meta-analysis p-value < 5×10^−8^. Those genome-wide significant SNPs identified in our EA population were then tested in the AA study population. Statistical significance for these SNPs was defined as a p-value <0.05/(number of significant SNPs in EA). We also compared our GWAS results to previously identified loci in other studies including SNPs in *ANXA11* (rs 1049550, rs1953600, rs2573346, rs2784773)[17], *RAB23* (rs1040461)[18], *C100RF67* (rs1398024)[19], *OS9* (rs1050045)[20], *CCDC88B* (rs479777)[21], and *NOTCH4* (rs715299)[4]. Statistical significance for these *a priori* SNPs was defined as a p-value<0.0056 (0.05/9 SNPs).

*Classic HLA alleles analysis*

We used logistic regression models to test for the association between the dosage of each imputed HLA allele and sarcoidosis susceptibility. Given the strong *a priori* nature of the HLA region, we used a p-value < 0.00011 (0.05/448 HLA alleles tested) to define statistical significance. We also compared our HLA alleles analysis results to previously identified HLA alleles in other studies, including DRB1*1101, DRB1*1501, and DQB1*0602[22, 23]. We used a p-value threshold of 0.016 (0.05/3 alleles) to determine statistical significance for these alleles chosen *a priori*.

*Conditional Models*

To assess the independence of single-SNP effects from the HLA risk alleles, we computed a multivariable logistic regression model where the HLA risk alleles were included as covariates in the model and each SNP, one at a time, was tested for association (i.e., association adjusted for HLA risk alleles).

*Expression quantitative locus (eQTL) and colocalization analysis*

For the subset of sarcoidosis cases with gene expression data available through GRADS, we performed colocalization analysis using eCAVIAR[24] to identify variants with evidence for colocalization of disease and *cis* eQTL associations. Here, the algorithm estimates the posterior probability that the same variant is casual in both the GWAS and eQTL studies while accounting for linkage disequilibrium (LD); we assumed one or two causal SNPs per region consistent with our results. We first identified the gene of the significant SNPs by searching the nearest gene within 25K of that SNP. The gene boundary was defined as 25K bases upstream/downstream of 5' and 3' UTR of the gene region defined by Genome Reference Consortium Human Build 37 (GRCh37) (https://www.ncbi.nlm.nih.gov/assembly/GCF_000001405.13/). We included all SNPs within the defined gene boundary of the significant SNP in the analysis. We then tested for association between those SNPs and gene expression. To provide the correlation matrix among the markers included in the eCAVIAR analysis, we used PLINK[25] to calculate the *r^2^* LD measure. The threshold for significance was set as a colocalization posterior probability (CLPP) >0.001, as suggested by the eCAVIAR developers[24]. The same approach was applied to the imputed HLA alleles. We tested for association between those HLA alleles and gene expression in two tissues (BAL and PBMC). We used Pearson's correlation matrix among those HLA alleles for the colocalization analysis. In addition to these analyses using the GRADS data, we also conducted a comprehensive cis-eQTL search using the publicly available database, Genotype-Tissue Expression (GTEx)[26, 27]. The GTEx project was supported by the Common Fund of the Office of the Director of the National Institutes of Health and by NCI, NHGRI, NHLBI, NIDA, NIMH, and NINDS. The data used for the analyses described in this manuscript were obtained from the GTEx Portal on 05/31/2020. The tissues included in the search were whole blood and lung biopsy (Inferior segment of left upper lobe, 1 cm below the pleural surface).

5.	Taliun D, Harris DN, Kessler MD, Carlson J, Szpiech ZA, Torres R, Taliun SAG, Corvelo A, Gogarten SM, Kang HM, Pitsillides AN, LeFaive J, Lee SB, Tian X, Browning BL, Das S, Emde AK, Clarke WE, Loesch DP, Shetty AC, Blackwell TW, Smith AV, Wong Q, Liu X, Conomos MP, Bobo DM, Aguet F, Albert C, Alonso A, Ardlie KG, Arking DE, Aslibekyan S, Auer PL, Barnard J, Barr RG, Barwick L, Becker LC, Beer RL, Benjamin EJ, Bielak LF, Blangero J, Boehnke M, Bowden DW, Brody JA, Burchard EG, Cade BE, Casella JF, Chalazan B, Chasman DI, Chen YI, Cho MH, Choi SH, Chung MK, Clish CB, Correa A, Curran JE, Custer B, Darbar D, Daya M, de Andrade M, DeMeo DL, Dutcher SK, Ellinor PT, Emery LS, Eng C, Fatkin D, Fingerlin T, Forer L, Fornage M, Franceschini N, Fuchsberger C, Fullerton SM, Germer S, Gladwin MT, Gottlieb DJ, Guo X, Hall ME, He J, Heard-Costa NL, Heckbert SR, Irvin MR, Johnsen JM, Johnson AD, Kaplan R, Kardia SLR, Kelly T, Kelly S, Kenny EE, Kiel DP, Klemmer R, Konkle BA, Kooperberg C, Kottgen A, Lange LA, Lasky-Su J, Levy D, Lin X, Lin KH, Liu C, Loos RJF, Garman L, Gerszten R, Lubitz SA, Lunetta KL, Mak ACY, Manichaikul A, Manning AK, Mathias RA, McManus DD, McGarvey ST, Meigs JB, Meyers DA, Mikulla JL, Minear MA, Mitchell BD, Mohanty S, Montasser ME, Montgomery C, Morrison AC, Murabito JM, Natale A, Natarajan P, Nelson SC, North KE, O'Connell JR, Palmer ND, Pankratz N, Peloso GM, Peyser PA, Pleiness J, Post WS, Psaty BM, Rao DC, Redline S, Reiner AP, Roden D, Rotter JI, Ruczinski I, Sarnowski C, Schoenherr S, Schwartz DA, Seo JS, Seshadri S, Sheehan VA, Sheu WH, Shoemaker MB, Smith NL, Smith JA, Sotoodehnia N, Stilp AM, Tang W, Taylor KD, Telen M, Thornton TA, Tracy RP, Van Den Berg DJ, Vasan RS, Viaud-Martinez KA, Vrieze S, Weeks DE, Weir BS, Weiss ST, Weng LC, Willer CJ, Zhang Y, Zhao X, Arnett DK, Ashley-Koch AE, Barnes KC, Boerwinkle E, Gabriel S, Gibbs R, Rice KM, Rich SS, Silverman EK, Qasba P, Gan W, Consortium NT-OfPM, Papanicolaou GJ, Nickerson DA, Browning SR, Zody MC, Zollner S, Wilson JG, Cupples LA, Laurie CC, Jaquish CE, Hernandez RD, O'Connor TD, Abecasis GR. Sequencing of 53,831 diverse genomes from the NHLBI TOPMed Program. *Nature* 2021: 590(7845): 290-299.

6.	Genomes Project C, Auton A, Brooks LD, Durbin RM, Garrison EP, Kang HM, Korbel JO, Marchini JL, McCarthy S, McVean GA, Abecasis GR. A global reference for human genetic variation. *Nature* 2015: 526(7571): 68-74.

7.	Karatzoglou A, Smola A, Hornik K, Zeileis A. kernlab - An S4 Package for Kernel Methods in R. *Journal of Statistic Software* 2004.

14.	Fingerlin TE, Zhang W, Yang IV, Ainsworth HC, Russell PH, Blumhagen RZ, Schwarz MI, Brown KK, Steele MP, Loyd JE, Cosgrove GP, Lynch DA, Groshong S, Collard HR, Wolters PJ, Bradford WZ, Kossen K, Seiwert SD, du Bois RM, Garcia CK, Devine MS, Gudmundsson G, Isaksson HJ, Kaminski N, Zhang Y, Gibson KF, Lancaster LH, Maher TM, Molyneaux PL, Wells AU, Moffatt MF, Selman M, Pardo A, Kim DS, Crapo JD, Make BJ, Regan EA, Walek DS, Daniel JJ, Kamatani Y, Zelenika D, Murphy E, Smith K, McKean D, Pedersen BS, Talbert J, Powers J, Markin CR, Beckman KB, Lathrop M, Freed B, Langefeld CD, Schwartz DA. Genome-wide imputation study identifies novel HLA locus for pulmonary fibrosis and potential role for auto-immunity in fibrotic idiopathic interstitial pneumonia. *BMC Genet* 2016: 17(1): 74.

15.	Fingerlin TE, Murphy E, Zhang W, Peljto AL, Brown KK, Steele MP, Loyd JE, Cosgrove GP, Lynch D, Groshong S, Collard HR, Wolters PJ, Bradford WZ, Kossen K, Seiwert SD, du Bois RM, Garcia CK, Devine MS, Gudmundsson G, Isaksson HJ, Kaminski N, Zhang Y, Gibson KF, Lancaster LH, Cogan JD, Mason WR, Maher TM, Molyneaux PL, Wells AU, Moffatt MF, Selman M, Pardo A, Kim DS, Crapo JD, Make BJ, Regan EA, Walek DS, Daniel JJ, Kamatani Y, Zelenika D, Smith K, McKean D, Pedersen BS, Talbert J, Kidd RN, Markin CR, Beckman KB, Lathrop M, Schwarz MI, Schwartz DA. Genome-wide association study identifies multiple susceptibility loci for pulmonary fibrosis. *Nat Genet* 2013: 45(6): 613-620.
